# Psilocybin modulation of dynamic functional connectivity is associated with plasma psilocin and subjective effects

**DOI:** 10.1101/2021.12.17.21267992

**Authors:** Anders S. Olsen, Anders Lykkebo-Valløe, Brice Ozenne, Martin K. Madsen, Dea S. Stenbæk, Sophia Armand, Morten Mørup, Melanie Ganz, Gitte M. Knudsen, Patrick M. Fisher

## Abstract

**Background:** Psilocin, the neuroactive metabolite of psilocybin, is a serotonergic psychedelic that induces an acute altered state of consciousness, evokes lasting changes in mood and personality in healthy individuals, and has potential as an antidepressant treatment. Examining the acute effects of psilocin on resting-state dynamic functional connectivity implicates network-level connectivity motifs that may underlie acute and lasting behavioral and clinical effects.

**Aim:** Evaluate the association between resting-state dynamic functional connectivity (dFC) characteristics and plasma psilocin level (PPL) and subjective drug intensity (SDI) before and right after intake of a psychedelic dose of psilocybin in healthy humans.

**Methods:** Fifteen healthy individuals completed the study. Before and at multiple time points after psilocybin intake, we acquired 10-minute resting-state blood-oxygen-level-dependent functional magnetic resonance imaging scans. Leading Eigenvector Dynamics Analysis (LEiDA) and diametrical clustering were applied to estimate discrete, sequentially active brain states. We evaluated associations between the fractional occurrence of brain states during a scan session and PPL and SDI using linear mixed-effects models. We examined associations between brain state dwell time and PPL and SDI using frailty Cox proportional hazards survival analysis.

**Results:** Fractional occurrences for two brain states characterized by lateral frontoparietal and medial fronto-parietal-cingulate coherence were statistically significantly negatively associated with PPL and SDI. Dwell time for these brain states was negatively associated with SDI and, to a lesser extent, PPL. Conversely, fractional occurrence and dwell time of a fully connected brain state was positively associated with PPL and SDI.

**Conclusion:** Our findings suggest that the acute perceptual psychedelic effects induced by psilocybin may stem from drug-level associated decreases in the occurrence and duration of lateral and medial frontoparietal connectivity motifs in exchange for increases in a uniform connectivity structure. We apply and argue for a modified approach to modeling eigenvectors produced by LEiDA that more fully acknowledges their underlying structure. Together these findings contribute to a more comprehensive neurobiological framework underlying acute effects of serotonergic psychedelics.

**Highlights:**

- We examined psilocybin effects on resting-state fMRI dynamic functional connectivity
- Diametrical clustering described as improved strategy for LEiDA-defined brain states
- Individual brain state dynamics defined by fractional occurrence and dwell time
- Two frontoparietal states and a fully connected brain state affected by psilocybin
- Brain state dynamics associated with psilocin level and subjective experience

## 1 Introduction

Psilocybin is a psychedelic compound that has gained significant interest over the last decade with promising evidence for therapeutic efficacy in treating several neurological and neuropsychiatric disorders, including depression (Carhart-Harris et al., 2021, 2018; Davis et al., 2021), anxiety (Vargas et al., 2020), substance abuse (Bogenschutz et al., 2015; Garcia-Romeu et al., 2019), migraine (Schindler et al., 2021), and cluster headache (Sewell et al., 2006). Through stimulation of the serotonin 2A receptor (5-HT2AR), psilocin, the neuroactive metabolite of psilocybin, potently and acutely induces an altered state of consciousness (Griffiths et al., 2006; Hasler et al., 2004; Madsen et al., 2019; Stenbæk et al., 2021). Psilocybin also induces rapid and lasting positive effects on mood, well-being, and personality (Carhart-Harris et al., 2018; Erritzoe et al., 2018; MacLean et al., 2011; Madsen et al., 2020). These intriguing effects precipitate the need to resolve associated and perhaps mediating neurobiological mechanisms. Such information can potentially inform future drug development programs and identify patient subgroups that may benefit from psychedelic therapy or predict potential adverse drug effects.

Previous studies have characterized distributed functional brain connectivity and macroscale cerebral networks acutely affected by a single administration of a serotonin psychedelic compound such as psilocybin with resting-state functional magnetic resonance imaging (rs-fMRI) (McCulloch et al., 2021a; Vollenweider and Preller, 2020). Studies suggest modulation of distributed connectivity patterns include alterations in thalamic connectivity (Preller et al., 2019), whole-brain connectivity (Madsen et al., 2021; Preller et al., 2020), decreased segregation and integration of canonical resting-state networks (Madsen et al., 2021; Mason et al., 2020), and macroscopic measures such as entropy (Roseman et al., 2014). However, most studies have focused on “static” functional connectivity, estimated as the correlation between pairs or across sets of areas over the duration of the scan session. This approach assumes signal stationarity for the entirety of the 5-10-minute rs-fMRI scan session, which may neglect relevant and observable neural dynamics arising from, e.g., mind-wandering or ephemeral experiences.

Dynamic functional connectivity (dFC) has emerged as a method for extracting informative, time-varying brain connectivity patterns (Preti et al., 2017). Unsupervised machine learning methods are employed to cluster instantaneous or small time-window connectivity metrics into discrete groups of distinct coactivation (Allen et al., 2014; Calhoun et al., 2014; Deco and Kringelbach, 2016). Such metrics are usually model-based and include lagged and zero-lag correlation coefficients and various estimates of interregional functional synchrony (Bastos and Schoffelen, 2016; Glerean et al., 2012). An appealing aspect of dFC strategies is that they attempt to model dynamics of connectivity processes that occur within a resting-state scan session, which is particularly relevant to the evaluation of psychedelics, which induce a dynamically evolving psychological experience.

Acute psychedelic effects on dynamic functional brain connectivity have been previously examined in only two separate datasets (Lord et al., 2019; Luppi et al., 2021; Tagliazucchi et al., 2014). Lord and colleagues applied Leading Eigenvector Dynamics Analysis (LEiDA) (Cabral et al., 2017) to rs-fMRI data acquired before and after a single intravenous dose of psilocybin in nine subjects. The authors reported that the probability of occurrence (“fractional occurrence”) of a discrete brain state comprising frontoparietal network elements was significantly lower after psilocybin infusion (Lord et al., 2019). Notably, plasma psilocin levels were not measured, which we have shown is tightly coupled to 5-HT2AR drug occupancy (Madsen et al., 2019) at the time of functional brain imaging. Moreover, there was only partial agreement between the discrete brain states identified and canonical resting-state networks, suggesting that acute psilocin effects may be informatively characterized by approaches that group sets of regions in a data-driven manner (e.g., clustering) as opposed to a priori defined network structures. It is critical to evaluate whether similar findings are observed in an independent sample and to evaluate this effect following oral psilocybin administration, as this is how it is administered clinically. During oral administration, the psychedelic effects are protracted over approximately six hours. Examining psilocybin effects on functional connectivity in alignment with an assessment of plasma psilocin levels and subjective effects throughout this period provides a novel perspective on its dynamic effects on the brain.

Furthermore, we see an opportunity to improve LEiDA and associated statistical evaluations. Typically, LEiDA clusters leading eigenvectors of instantaneous phase coherence maps using Euclidean k-means. Prior to clustering, eigenvectors are flipped so that the majority of elements are negative. However, eigenvectors are, in practice, normalized to have unit length and have arbitrary sign. Thus, eigenvectors are distributed on the antipodally symmetric unit hypersphere, attributes not acknowledged by Euclidean k-means nor preserved by the aforementioned flip procedure (see Supplementary Figure S1). This leads to sub-optimal clustering. Directional statistics is a branch of statistics that deals with data where the direction holds more information than the amplitude, typically represented as normalized vectors distributed on some geometric manifold (Mardia and Jupp, 1999). Specifically, the Watson distribution (Watson, 1965) is optimal for modeling data distributed on the antipodally symmetric unit hypersphere. Diametrical clustering (Dhillon et al., 2003; Sra and Karp, 2013) is the k-means equivalent of Watson mixture models and may offer more suitable clustering of eigenvectors in LEiDA. In addition to fractional occurrence, the average duration of brain state occurrences (“dwell time”) can complementarily inform the nature of connectivity dynamics. We suggest modeling dwell time using survival analysis, which more appropriately captures the conditional dependence of state probability on the previous length of active time (Cox, 1972).

Here we evaluated acute psilocybin effects on dFC with blood-oxygen-level-dependent (BOLD) rs-fMRI in 15 healthy participants, each of whom completed one 10-min rs-fMRI scan session before intake of a psychedelic dose of psilocybin and multiple 10-min rs-fMRI scan sessions after psilocybin intake (approximately 40, 80, 140 and 300-min post-administration). We applied LEiDA with diametrical clustering to account for the intrinsic spherical geometry and antipodal symmetry in the distribution of eigenvectors. To establish the association between dFC characteristics and the psychopharmacological effects of psilocybin, we determined the association between the fractional occurrence of discrete brain states, defined by clustering, and both plasma psilocin level (PPL) and subjective drug intensity (SDI), which we have shown are coupled to 5-HT2AR occupancy and baseline 5-HT2AR (Madsen et al., 2021, 2019; Stenbæk et al., 2021). We determined associations between PPL and SDI and discrete brain state dwell time using Cox regression frailty models.

## 2 Methods

A brief description of experimental procedures is provided here; a full description can be found elsewhere (Madsen et al., 2021).

### 2.1 Experimental procedures

Fifteen healthy participants (age 34.3 ± 9.8 years, six females) with no or limited prior experience with psychedelics were recruited for a brain imaging study, including a single psilocybin intervention. All participants provided written informed consent and were healthy, including screening for neurological, psychiatric, or somatic illnesses. Two psychologists prepared participants and supported them at all stages of the intervention.

Psilocybin was taken orally in multiples of 3 mg psilocybin capsules, dosed according to body weight (total dose: 0.24 ± 0.04 mg/kg) in a single-blind cross-over study design. Within each cross-over, participants received either psilocybin or a non-psychedelic drug (ketanserin); only the psilocybin data are reported here. Functional neuroimaging data were acquired once before and at regular intervals (approximately 40, 80, 130, and 300 minutes) after administration. Immediately after each rs-fMRI acquisition, participants were asked to rate their perceived SDI on a Likert scale from 0 to 10 (0 = “not at all intense”, 10 = “very intense”). Following each subjective rating, a blood sample was drawn from an intravenous catheter to determine the concentration of unconjugated psilocin in plasma (Kolaczynska et al., 2021; Madsen et al., 2021). The study was approved by the ethics committee for the capital region of Copenhagen (journal identifier: H-16,028,698, amendments: 56,023, 56,967, 57,974, 59,673, 60,437, 62,255) and the Danish Medicines Agency (EudraCT identifier: 2016–004,000– 61, amendments: 2,017,014,166, 2,017,082,837, 2,018,023,295).

### 2.2 Neuroimaging data acquisition

MRI data were acquired on a 3T Siemens Prisma scanner (Siemens, Erlangen, Germany) with a 64-channel head coil. A structural T1-weighted 3D image was acquired at the pre-drug imaging session (inversion time = 900 ms, TE/TR = 2.58/1900 ms, flip angle = 9°, matrix 256×256×224, resolution 0.9 mm isotropic, no gap). BOLD fMRI data were acquired using a T2*-weighted gradient echo-planar imaging sequence (TE/TR = 30/2000 ms, flip angle = 90°, in-plane matrix = 64×64 mm, in-plane resolution = 3.6×3.6 mm, 32 slices, slice thickness = 3.0 mm, gap = 0.75 mm). 300 volumes (10 minutes) were acquired in each imaging session. Participants were instructed to close their eyes and let the mind wander freely without falling asleep. In total, 74 scan sessions were acquired across the 15 participants.

### 2.3 fMRI data preprocessing

fMRI data preprocessing was performed separately for each of the 10-minute rs-fMRI scan sessions. The data were preprocessed in SPM12 (http://www.fil.ion.ucl.ac.uk/spm). Steps included 1) slice-timing correction, 2) spatial realignment and field unwarping, 3) co-registration of the T1-weighted structural image to the first functional volume, 4) segmentation of the T1-weighted image into gray matter, white matter, and cerebrospinal fluid (CSF) maps, 5) normalization of the Anatomical Automatic Labeling (AAL) atlas (Tzourio-Mazoyer et al., 2002) to the co-registered structural images, and 6) smoothing of functional images (4mm FWHM Gaussian kernel). Motion and signal variance artifacts were identified using Artifact detection Tool (ART, https://www.nitrc.org/projects/artifact_detect). Individual scan sessions where more than 50% of volumes exceeded the ART threshold were excluded from the analysis. Based on this criterion, two scan sessions from a single participant were excluded, resulting in 21,600 rs-fMRI volumes included in subsequent analyses. fMRI time-series were denoised using CONN (Whitfield-Gabrieli and Nieto-Castanon, 2012) by voxel-wise nuisance regression of 1) three translation and three rotation parameters from realignment and their first-order derivatives, and 2) anatomical component correction using the first five principal components and their first-order derivatives from white-matter and CSF time-series (Behzadi et al., 2007). Time-series data were bandpass filtered between 0.008 and 0.09 Hz. We used the AAL atlas to parcellate the denoised functional images into 90 cortical and subcortical regions.

### 2.4 Extraction of BOLD phase-series

For each scan session, we estimated regional phase-series from the BOLD signals *S*(*t*) by constructing the analytic signal *Z*(*t*) = *S*(*t*) + *iS*_*h*_ (*t*), where *i* is the imaginary unit and 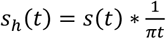 is the Hilbert transform, where * represents the convolution operator (see Figure 1). The analytic signal is circularly evolving and can thus be described by instantaneous (i.e., per time point) amplitude 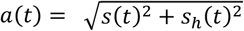 and phase 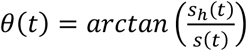. The instantaneous phase is a sawtooth curve representing the locally linear temporal phase angle variation with a discontinuity at the jump from −*π* to *π* (see Figure 1A, 2^nd^ panel), and generally contains the oscillatory information in *S*(*t*), while *a*(*t*) encompasses (potentially spurious) amplitude information. Instantaneous phase coherence between brain region pairs (*j, k*) ∈ {1, …, 90}^2^ was described using the symmetric phase coherence map ***A***_*t*_ for every time point *t* with elements *A*_*t,j,k*_ = *cos*(*θ*_*t,j*_ − *θ*_*n,k*_). Since ***A***_*t*_ has 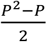 unique elements, we may be well served by describing its information in lower dimensions using the eigendecomposition. Due to the angle difference identity *cos*(*θ*_*j*_ − *θ*_*k*_) = *cos θ*_*j*_ *cos θ*_*k*_ + Sin *θ*_*j*_ Sin *θ*_*k*_, for any two regions *j* and *k*, ***A*_*t*_** can be fully decomposed into two P-dimensional orthogonal eigenvectors, which are each a linear combination of the vectors ***c*** = *cos* ***θ***_*t*_ and ***s*** = Sin *θ*_*t*_. For each time point, we retained only the eigenvector ***v***_1,*t*_ corresponding to the largest eigenvalue, thereby capturing the dominant instantaneous connectivity pattern.

**Figure 1:**
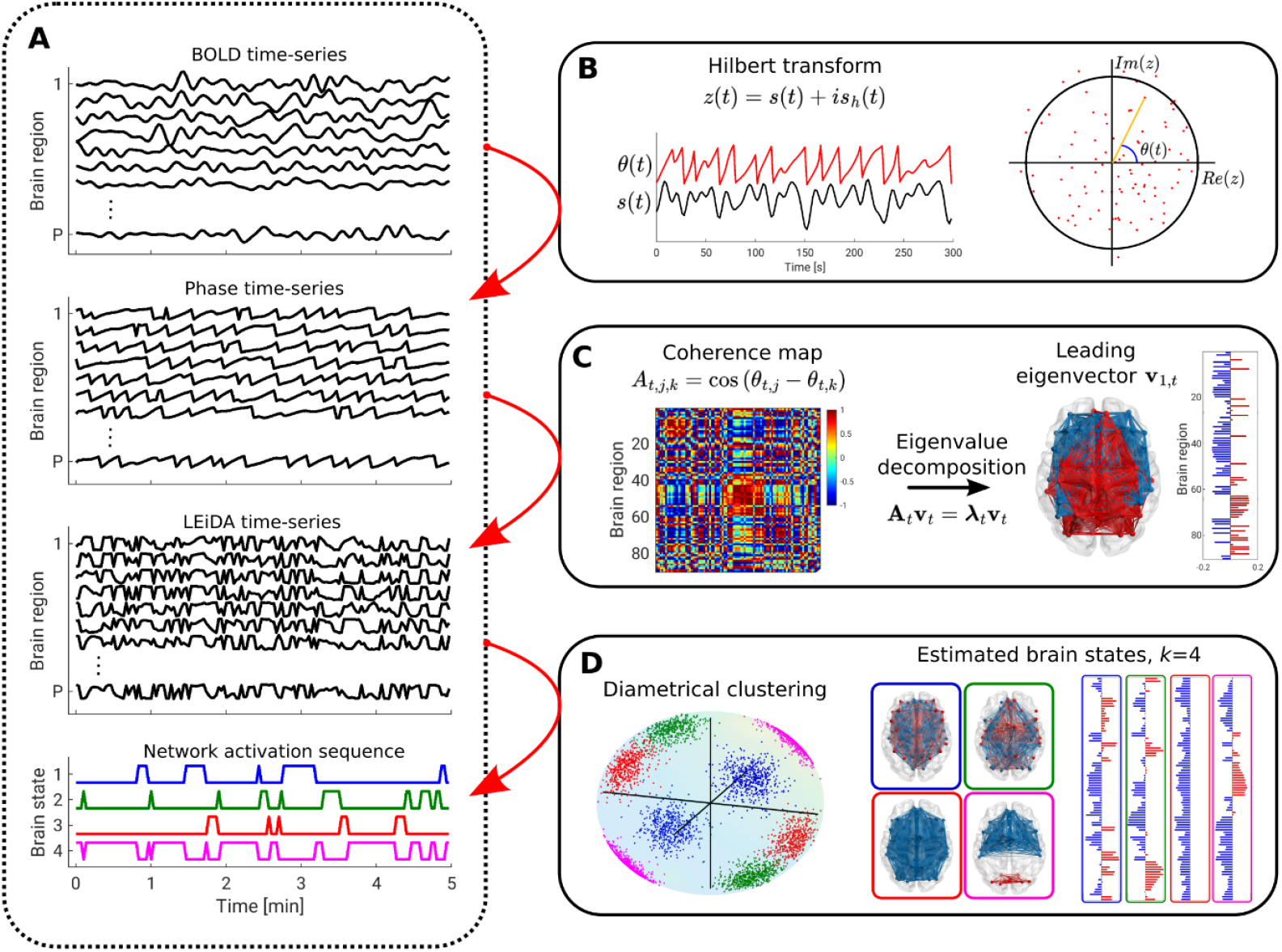
Methodological pipeline for leading eigenvector dynamics analysis (LEiDA) and diametrical clustering. (A): The LEiDA pipeline consists of extraction of session and region-wise instantaneous BOLD phases via the Hilbert transform (B), followed by an eigenvalue decomposition of the associated phase coherence map for every time-point, t (C). The eigenvectors are constrained to unit norm. Diametrical clustering is applied to the set of leading eigenvectors across all scan sessions and subjects to derive discrete brain states (D). Every volume is hard-assigned to the closest brain state, generating the network activation sequence (A, lower panel), after which session-specific brain state descriptors such as fractional occurrence or dwell time may be calculated.

### 2.5 Clustering

Eigenvectors are usually normalized to unit length and have arbitrary sign, and are thus distributed on the surface of the antipodally symmetric unit hypersphere. The (Dimroth-Scheidegger)-Watson distribution models such data (Sra and Karp, 2013; Watson, 1965). If we assume that eigenvectors can be described by a mixture of independent Watson distributions, we can disentangle clusters by estimating a Watson mixture model. Diametrical clustering is derived from mixture modeling of multivariate Watson distributions where only the mean direction is modeled, disregarding cluster variance and covariance structures (Dhillon et al., 2003; Sra and Karp, 2013). Diametrical clustering can be regarded as the standard k-means algorithm where centroid locations are updated according to the squared Pearson correlation similarity measure: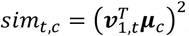, where ***µ***_*c*_ is the centroid of cluster *c*, and ***v***_1,*t*_ the leading eigenvector of the phase coherence map for any time point *t* or scan session. Diametrical clustering may be described as the limit where all Watson distributions in the mixture have shared concentration parameter *k* → ∞. Importantly, this approach addresses two limitations of previously applied clustering techniques, namely 1) the ability to group correlated and anticorrelated unit norm vectors into the same cluster and 2) by constraining the optimization to the surface of the associated hypersphere. The squared Pearson correlation is equivalent to the squared cosine similarity for normalized vectors, and thus equivalent to finding the squared cosine of the angle between the unit vectors. We initialized our algorithm using k-means++ rewritten for diametrical clustering (Arthur and Vassilvitskii, 2007). For each *k*, 5 replications of the clustering algorithm were run, where the best of the 5 replications (in terms of the sum of squared Pearson correlation to the nearest centroid) was chosen as the output.

To retrieve recurrent interregional phase coherence patterns, we grouped the 21,600 leading eigenvectors into *k* clusters, which we denote “brain states” (see Figure 1D). The optimal number of brain states is not known or well-defined; therefore, we produced models for *k* ranging from 2 to 20, with higher *k* revealing more fine-grained patterns (Cabral et al., 2017; Figueroa et al., 2019; ML et al., 2020)¶.

### 2.6 Identification of recurrent brain states

The diametrical clustering algorithm returns *k* unordered cluster directions and labels of volumes assigned to each model according to the squared Pearson correlation. To match brain states across *k*, the estimated centroids for *k* were assigned to the closest centroid for *k* − 1, without replacement. To match specific centroids across different *k*, we selected a template *k*, and, for all other *k*, found the centroid that most closely matched the template centroid in terms of squared Pearson correlation. To analyze clustering stability, we ran diametrical clustering 1000 times with five replications each for all *k* ∈ {2, …, 20}, and identified, for each *k*, the state most closely matching the relevant template states. If the Pearson correlation coefficient between the identified states and the template was negative, the sign of the identified state was inverted. We also compared diametrical clustering output to that of Euclidean k-means, where the output centroids from the latter were normalized to unit length before matching.

### 2.7 Brain network state occurrence

To identify associations between brain state dynamics and PPL and SDI, we calculated the fractional occurrence for each brain state, as defined by the fraction of time points in a scan session assigned to that brain state. For each state, this produced 72 FO estimates, one for each scan session. We modeled the association between FO and PPL (or SDI) with a random intercept linear mixed-effects model to account for inter-subject variability. The models were fitted using maximum likelihood, and we used the likelihood ratio to test for significance of the fixed effect and generate confidence intervals unadjusted for multiple comparisons (CI_unadj_). To account for multiple testing across a set of *k* states, we performed permutation testing with max-T adjustment and 100,000 permutations by scrambling the normalized residuals of the linear mixed-effects model (Lee and Braun, 2012; Westfall and Young, 1993). The initially observed statistical estimates (likelihood ratios) were then compared to the distribution of maximum statistics across the *k* models for every permutation, and a corresponding p-value was calculated as the number of permutations where the initial likelihood ratio exceeded the maximum statistic. Permutation testing and Max-T correction were performed within-*k* and separately for the models with PPL and SDI as fixed effects, respectively.

### 2.8 Brain network state dwell time

We employed survival analysis to model state dwell time, i.e., the time spent in a brain state before switching. Whenever the brain state changed within a scan session, we noted the number of preceding samples *t* and the corresponding subject, PPL, and SDI. The first active state from a scan session was excluded since we cannot estimate the true dwell time in this case. We performed right censoring for the last active state in a scan session. We modeled the dwell time of a brain state using a Cox proportional hazards model, including a frailty element *z* to account for inter-subject variability: *λ*(*t*|*x*_*nl*_, *Z*_*n*_) = *Z*_*n*_*λ*_0_(*t*) exp(*x*_*nl*_ *β*), where *n* = 1, …, *N* denotes subjects *l* = 1, …, *N*_*n*_ denotes sessions for subject *n*, and *x*_*nl*_ the corresponding PPL or SDI (Cox, 1972; Vaupel et al., 1979). We report the estimated hazard ratio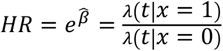, which is proportional in the covariate level (PPL or SDI). The associated confidence interval is defined as *e*^*β*±1.96*se*(*β*)^, where *Se*(*β*) is the estimated standard error of the coefficient estimate. *λ*_0_(*t*) represents the common baseline hazard irrespective of subject or covariate level. We could not find any existing permutation test specifically for frailty Cox proportional hazards models. Therefore, we controlled the FWER using Bonferroni-Holm correction (Holm, 1979) applied within-*k*.

### 2.9 Visualizations, code, and data availability

We have published the MATLAB (The MathWorks, inc.) and R code used to generate the results presented in this study at (https://github.com/anders-s-olsen/psilocybin_dynamic_FC). We used BrainNet Viewer(Xia et al., 2013) (https://www.nitrc.org/projects/bnv/) to generate connectivity visualizations. The datasets generated and/or analyzed during the current study can be made available upon completion of a formal data sharing agreement.

## 3 Results

21,600 leading eigenvectors defined by LEiDA were clusteres into *k* discrete brain states defined by centroids determined with diametrical clustering (see Figure 1). We explored a range of *k* ∈ {2, …, 20} centroids, aligned with previous studies (Cabral et al., 2017; Figueroa et al., 2019; Kringelbach et al., 2020). Generally, specific centroid locations were stable across contiguous values of *k*; 90-dimensional projections of all centroids for all values of *k* can be found in Supplementary Video S2. See Figure 2 for a visualization of estimated brain states for *k* = 7.

**Figure 2:**
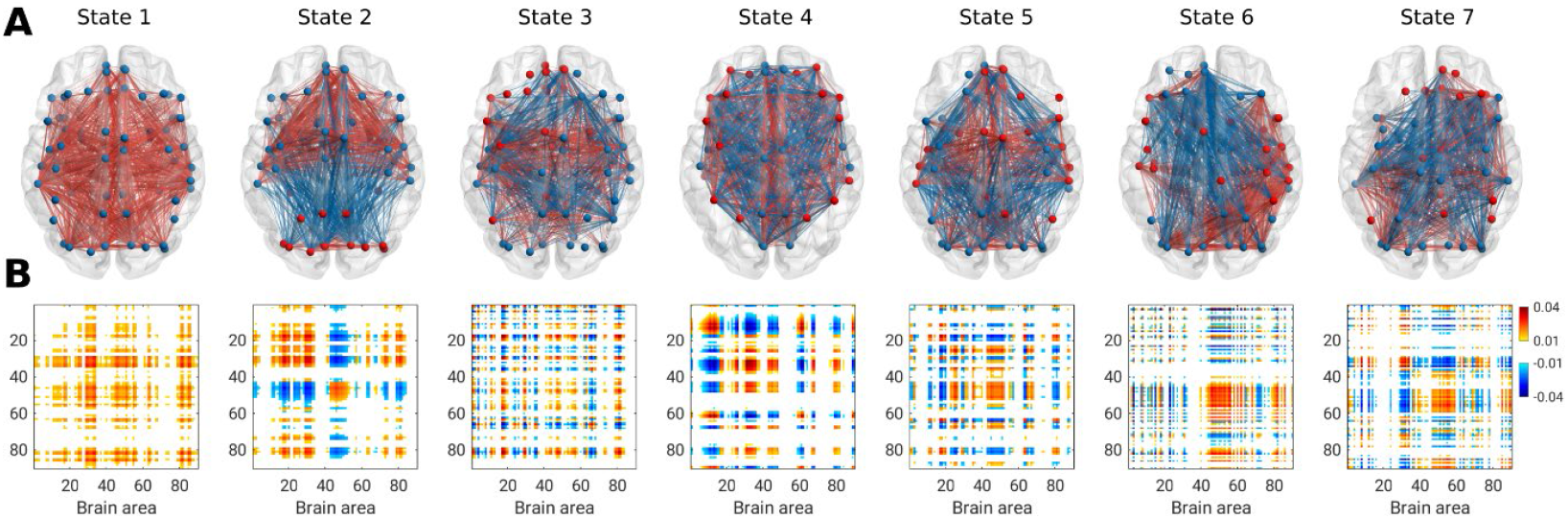
Brain states estimated using LEiDA and diametrical clustering with k=7 with spatial connectivity representation (A) and coherence maps defined as the outer product of the cluster centroid (B). Only edges above the 75^th^ percentile of absolute edge strengths are shown. In (A), all positive edges are shown in red and negative edges in blue, while nodes are colored according to the sign of their element in the respective cluster centroid.

For all values of *k* ≥ 4, we observed a “global” brain state characterized by all centroid elements having the same sign (e.g., see state 1 in Figure 2). All other brain states were characterized by coherence loadings in both directions. For example, for *k* = 7, brain state 2 showed strong coherence between areas related to visual processing (red nodes) and regions related to the salience network (blue nodes). Brain state 4 showed coherence between regions related to the frontoparietal, or central executive network, including the dorsolateral prefrontal cortex and posterior parietal cortex. Anti-coherent elements were observed in cingulate and parietal regions.

### 3.1 Psilocybin effects on brain state fractional occurrence and dwell time

Figures 3a and 3c summarize FWER-controlled p-values of linear mixed-effects models estimates of the association between fractional occurrence (FO) of individual brain states and PPL or SDI, respectively. Across multiple values of *k* ≥ 4, we observed one brain state (“frontoparietal state 1”, green triangle symbols in Figure 3, see also Figure 4) for which the FO was statistically significantly negatively associated with both PPL (*k = 7*: slope = −0.0066, 95% CI_unadj_ = [−0.0094;−0.0038]; p_FWER-maxT_ < 0.001; units: FO per μg/ml PPL; Figure 4; Supplementary Table S3) and SDI (*k = 7*: slope = −0.014, 95% CI_unadj_ = [−0.017;−0.010]; p_FWER-maxT_ < 0.001; units: FO per SDI rating; Figure 4; Supplementary Table S3). Specifically, the association between brain state FO and PPL was significant for the interval *k* ∈ {4, …, 9}, whereas for SDI, this association was significant for all *k* ≥ 4. Put another way, the average total time the brain occupies this frontoparietal state during the course of a 10-min rs-fMRI scan was negatively related to PPL and SDI. For *k* ≥ 8, we observed a second brain state (“frontoparietal state 2”, red star symbols in Figure 3, see also Supplementary Figure S4) for which the FO was also statistically significantly negatively associated with both PPL (*k = 11*: slope = −0.0039, 95% CI_unadj_ = [−0.0058;−0.0021]; p_FWER-maxT_ = 0.001; units: FO per μg/ml PPL; Supplementary Figure S4; Supplementary Table S3) and SDI (*k = 11*: slope = −0.0076, 95% CI_unadj_ = [−0.0101;−0.0050]; p_FWER-maxT_ < 0.001; units: FO per SDI rating; Supplementary Figure S4; Supplementary Table S3). For *k* ≥ 4, we observed a third brain state (“fully connected state”, blue diamond symbols in Figure 3, see also Supplementary Figure S5) for which the FO was statistically significantly positively associated with SDI (*k = 7*: slope = 0.0076, 95% CI_unadj_ = [0.0024;0.0127]; p_FWER-maxT_ = 0.035; units: FO per SDI rating; Supplementary Figure S5; Supplementary Table S3). We did not observe any statistically significant associations between the fully connected state and PPL.

**Figure 3:**
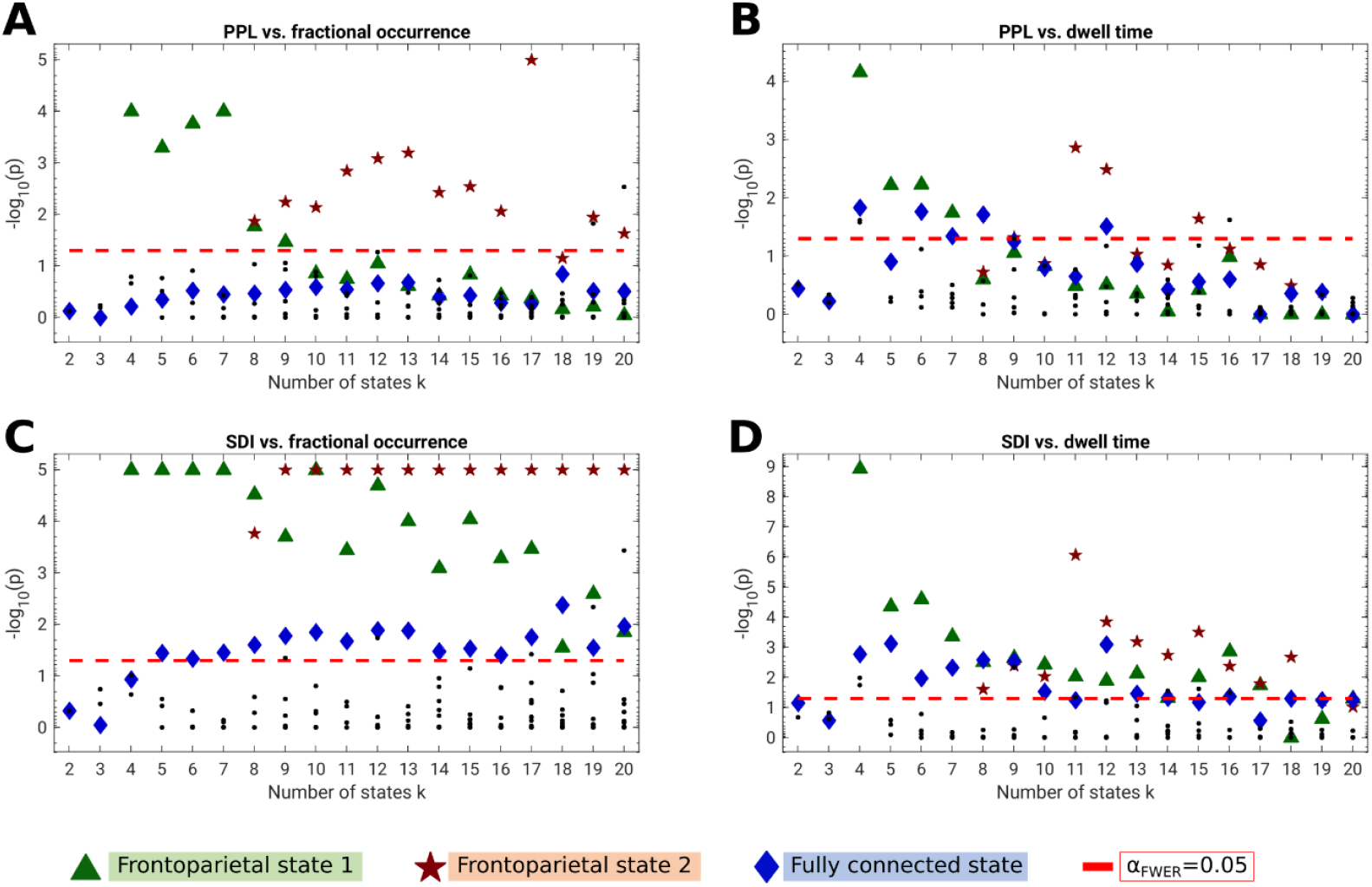
Summary statistics linking brain state fractional occurrence and dwell time with plasma psilocin level (PPL) and subjective drug intensity (SDI). (A): Linear mixed-effects models of the association between PPL and brain state fractional occurrence. (B): Cox proportional hazards frailty models of the association between PPL and brain state dwell time. (C): Linear mixed-effects models of the association between SDI and brain state fractional occurrence. (D): Cox proportional hazards frailty models of the association between SDI and brain state dwell time. Horizontal red line denotes family-wise error rate (FWER) threshold for statistical significance. Fractional occurrence p-values were corrected using 100,000 permutations and max-T correction applied within-k (see Methods). Where an observed statistic exceeded all permuted values, the p-value was set to 10^−5^ (i.e., − log_10_(p) = 5). Dwell time p-values were corrected using Bonferroni-Holm applied within-k. For every k, points were identified as one of the three brain states by matching all the corresponding centroids to the templates (k=7 for frontoparietal state 1 and the fully connected state, k=11 for frontoparietal state 2).

**Figure 4:**
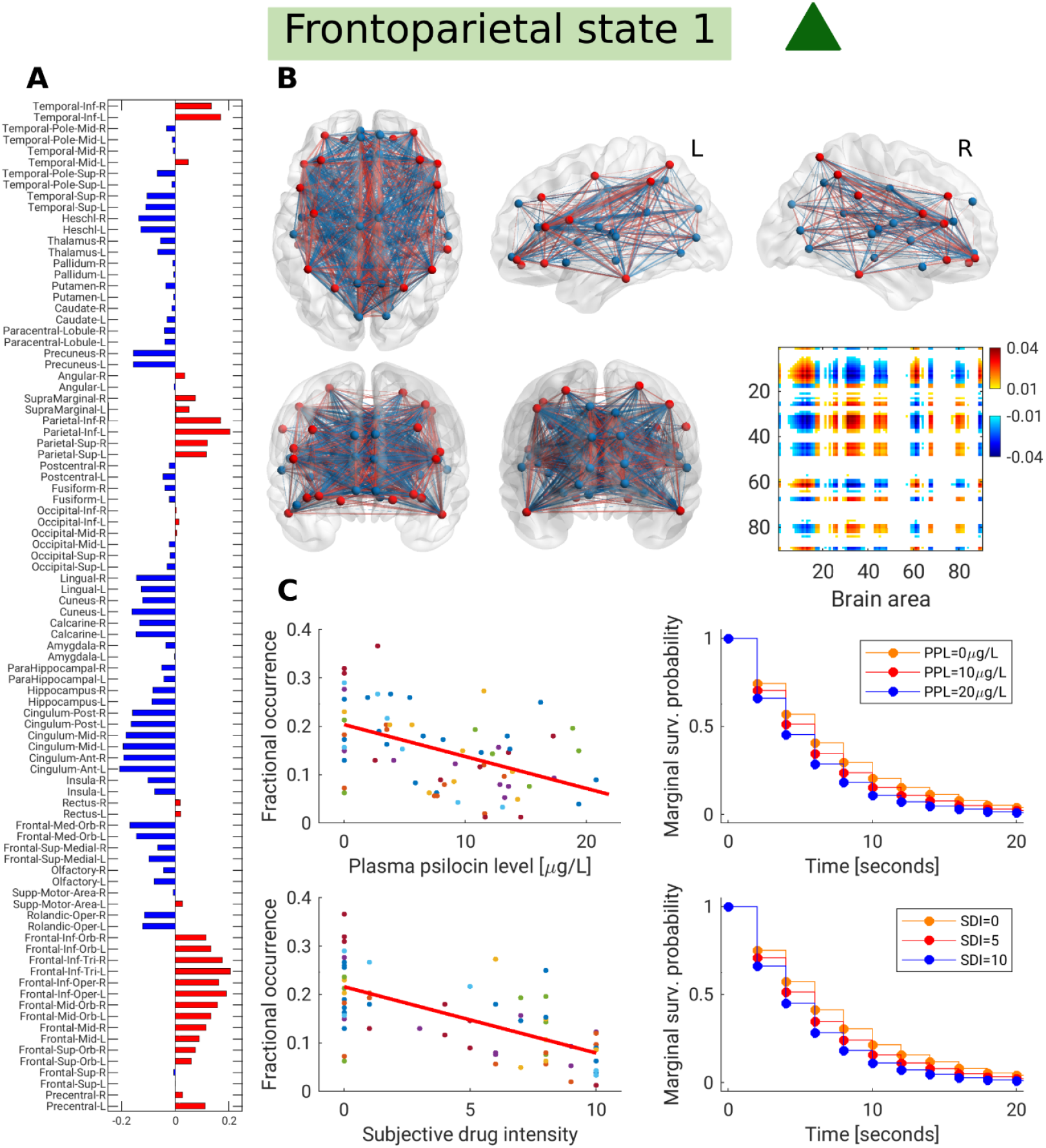
Frontoparietal state 1 and statistical associations for k=7. (A): 90-dimensional centroid, where region pairs with the same sign are said to be coherent. Many frontal regions, superior and inferior parietal regions, and inferior temporal lobe showed coherence with each other (red). Likewise, areas around the parieto-occipital sulcus, cingulum, and medial orbital frontal cortex were coherent (blue). (B): Functional coherence map and connectivity representation of the brain state. Edges are shown if their strength exceeds the 75^th^ percentile of absolute edge strengths. In the connectivity visualizations, negative edges are blue and positive edges are red, while nodes are colored according to the sign of their centroid element in (A). (C): Associations between the expression of frontoparietal state 1 and plasma psilocin level and subjective drug intensity using linear mixed-effects models for fractional occurrence (left, each point is a scan session) and frailty Cox proportional hazards models for dwell time (right). For dwell time, the marginal survival curves for pre-specified covariate levels are shown. Colors in plots of fractional occurrence (C, left) denote individual participants.

Figure 3b and 3d summarize Bonferroni-Holm corrected p-values of Cox proportional hazards models of the effect of PPL and SDI on dwell time, respectively. Frontoparietal state 1 dwell time was negatively associated with PPL across several values of *k* (*k* = *7*; Hazard ratio (HR) = 1.017, 95% CI_unadj_ = [1.006;1.028]; p_FWER-BH_ = 0.018; Figure 4; Supplementary Table S3) and SDI (*k* = *7*; HR = 1.037, 95% CI_unadj_ = [1.018;1.055]; p_FWER-BH_ < 0.001; Figure 4; Supplementary Table S3). Specifically, the association between brain state dwell time and PPL was statistically significant for *k* ∈ {4, …, 7}, whereas for SDI, this association was statistically significant for *k* ∈ {4, …, 17,20}. In other words, the higher the PPL and SDI, the larger the hazard ratio, i.e., the less average continuous time spent in frontoparietal state 1. For *k* ≥ 8, frontoparietal state 2 also showed a negative association with both PPL (*k* = *11*; HR = 1.027, 95% CI_unadj_ = [1.013;1.040]; p_FWER-BH_ = 0.001; Supplementary Figure S4; Supplementary Table S3) and SDI (*k* = *11*; HR = 1.057, 95% CI_unadj_ = [1.036;1.079]; p_FWER-BH_ < 0.001; Supplementary Figure S4; Supplementary Table S3). Overall, frontoparietal state 2 was significantly inversely associated to PPL for *k* = {9,11,12,15} and to SDI for *k* ∈ {8, …, 18}. For *k* ≥ 4 we observed that dwell time of the fully connected state was positively associated with both PPL (*k* = *7*; HR = 0.983, 95% CI_unadj_ = [0.972;0.996]; p_FWER-BH_ = 0.045; Supplementary Figure S5; Supplementary Table S3) for *k* = {4,6,7,8,12} and SDI (*k* = *7*; HR = 0.970, 95% CI_unadj_ = [0.952;0.987]; p_FWER-BH_ = 0.005; Supplementary Figure S5; Supplementary Table S3) for *k* ∈ {4, …, 10,12,13,14,16}.

All centroids across all values of *k* showing a statistically significant association between either PPL or SDI and either FO or dwell time are listed in Supplementary Table S3.

### 3.2 Stability of highlighted states

To identify the three brain states across *k*, we defined template centroids (*k* = 7 for frontoparietal state 1 and the fully connected state, *k* = 11 for frontoparietal state 2, see Supplementary Figures S6–8). For every *k*, the brain state most closely matching each of these three templates were marked. In Supplementary Figure S9, between-*k* correlation coefficients indicate that the three states had very similar centroids across the range of *k*. The between-*k* similarity can also be confirmed visually in Supplementary Figures S6–S8. Frontoparietal state 2 appeared initially at *k* = 8 and qualitatively became more associated with PPL and SDI than frontoparietal state 1 (see Figure 3). Likewise, estimated fractional occurrence slopes for the association between frontoparietal state 1 and PPL and SDI approximately halved at the transition from *k* = 7 to *k* = 8 (Supplementary Table S3), suggesting that frontoparietal state 2 was incorporated within frontoparietal state 1 for *k* < 8.

### 3.3 Diametrical clustering stability and comparison to k-means

Like most clustering algorithms, diametrical clustering is initialized randomly. To quantify the variation in brain state centroid location between initializations, we ran diametrical clustering 1000 times with five replications and extracted the two frontoparietal states and the fully connected state. Supplementary Figure S10 shows the histogram of Fisher’s r-to-z scores of the Pearson correlation coefficients across all initialization pairs, including a fitted Gaussian curve. Generally, we see high clustering stability regardless of initialization. The average correlation coefficient between initializations is numerically higher for the fully connected state, followed by frontoparietal states 1 and 2. As expected, stability decreases with increasing *k*.

We compared brain state-specific differences in centroid locations between those obtained using the diametrical clustering method presented here and Euclidean k-means used in previous LEiDA-studies. Notably, at *k* = 7, the “fully connected state” is not identified when using the Euclidean k-means approach for clustering (Supplementary Figure S11). Although there are clear similarities across the brain states paired between the two clustering methods, the magnitudes of these similarities are variable. To understand this variability more comprehensively, we ran both diametrical clustering and LEiDA Euclidean k-means without replications using 1000 initializations and computed the correlation coefficient for all 1000×1000 combinations between the two methods for each of the extracted centroids for frontoparietal state 1, frontoparietal state 2, and the fully connected state. The mean, *µ*_*ρ*_, and standard deviation, *σ*_*ρ*_, of these correlation coefficients are described in Supplementary Table S12. Although often highly correlated, these results show variability in the correlation between these two clustering methods, suggesting they do not always produce convergent results.

## 4 Discussion

Here we evaluated acute psilocybin effects on dynamic functional brain connectivity in healthy individuals. Most prominently, the higher the subjective experience intensity and plasma psilocin level, the lower the fractional occurrence of two discrete frontoparietal-like brain states. Similarly, the average dwell time of these brain states was inversely related to plasma psilocin level and subjective drug intensity. We observed an increase in the fractional occurrence and dwell time of a “fully-connected” brain state where all elements have the same sign, although the statistical associations for this state were weaker. Together, these findings provide a novel mapping of drug availability and perceptual intensity of a clinically relevant psilocybin-induced psychedelic experience onto distributed whole-brain functional connectivity dynamics. We propose an alternative method for clustering LEiDA-dFC estimates that we believe more faithfully respects the spherical manifold and sign ambiguity of orthonormal eigenvectors (Dhillon et al., 2003; Sra and Karp, 2013). Taken together, these findings implicate dynamic neural processes underlying the acute psychedelic effects of psilocybin, an important contribution to understanding the effects of this rapidly emerging clinical therapeutic.

The highlighted frontoparietal states 1 and 2 were both characterized by phase coherence between areas commonly assigned to a network described as, e.g., the “frontoparietal” network, “central executive”, “executive control”, or “dorsal attention” network (Witt et al., 2021). Similarly, these brain states expressed phase coherence between regions in the cingulum and some regions around the parieto-occipital fissure (see Figure 4 and Supplementary Figure S4). The regions with strong “negative” loadings were remarkably similar between the two states. The two states mostly differed in the centroid loadings for elements in the temporal lobe and the Rolandic operculum. A previous study applying LEiDA to model dynamic functional connectivity following psilocybin administration reported a similar brain state for models in the range *k* ∈ {5, …, 10} (Lord et al., 2019). Despite methodological differences between the studies, e.g., we administered psilocybin orally, measured PPL, scanned participants multiple times after administration, and applied diametrical clustering; it is encouraging that our findings offer convergent evidence that decreased frontoparietal connectivity is a critical neural characteristic of the psilocybin-induced drug experience. We show here, for the first time, that these changes are proportionally related to PPL and SDI across the duration of the psychedelic experience. Contributing to our mechanistic understanding of the neurobiological mechanisms that shape psilocybin effects, our findings implicate a systems-level neural correlate (frontoparietal state prevalence) to the relation between available psilocin, which we have previously shown to be associated with 5-HT2AR occupancy, and subjective intensity of the psychedelic experience (Madsen et al., 2019).

Consistent with the observed effects on fractional occurrence, we observed some evidence that dwell time, i.e., average time spent in the state before switching, of the frontoparietal states were similarly negatively associated with PPL and SDI. However, this effect was statistically significant for only a subset of the evaluated number of brain states, *k*. Notably, the integral of the subject-specific survival function for which hazard ratios were estimated is proportional to fractional occurrence. This means that dwell time models not merely the (instantaneous) probability of being in a given state but also the exponential decrease of that probability over consecutive time points. We infer that the numerically consistent associations with fractional occurrence and dwell time reflect a psilocybin-induced “bias shift” away from the observed frontoparietal brain states. Previous studies examining brain state switching mechanisms have typically evaluated transition probability matrices and specific state-to-state transition probabilities conditioned only on the current state. Although dwell time is related to the diagonal elements of the transition matrix, modeling it as a hazard ratio informs state survival across a broader time window, giving a more complete perspective on brain state dynamics. Modeling dwell time using survival analysis does not model all state-to-state transition probabilities individually. However, many of these transitions occur only rarely, and the set of statistical tests squares with the number of brain states, *k*, both of which constrain associated statistical estimates. In this way, we view the Cox proportional hazards model as a valuable trade-off for evaluating state dwell time and switching probability.

Previous studies applying LEiDA have reported alterations in a “fully connected” brain state, characterized by all elements having the same sign (Cabral et al., 2017; Escrichs et al., 2021; Farinha et al., 2021; Figueroa et al., 2019; Larabi et al., 2020; Lord et al., 2019; Stark et al., 2021; Vohryzek et al., 2020). Here we also observed this fully connected state and report an increase in fractional occurrence significantly associated with SDI, but not PPL. Interestingly, however, Supplementary Figure S11 shows that we would not have identified this fully connected state if we applied LEiDA using the Euclidean k-means clustering method described previously. The observed slope estimates for the fully connected state are similar, and opposite to those for the two frontoparietal states for *k* ≥ 8, and approximately half that of frontoparietal state 1 for *k* < 8. Similarly, dwell time for the fully connected state was significantly positively associated with both PPL and SDI. Here, hazard ratio estimates were similar for all three highlighted brain states regardless of *k*. These results indicate that while psilocin induces a decrease in the fractional occurrence and dwell time of frontoparietal connectivity dynamics, only approximately half of the corresponding increase in brain activity can be explained by a shift toward the fully connected state. As fractional occurrences must sum to one across all states, these findings suggest additional increases are spread across other states below the statistical significance threshold, given the current data.

Here we have presented the application of diametrical clustering, which we view as a fundamentally more appropriate clustering method than k-means based on Euclidean distance because eigenvectors are, in practice, normalized to unit length. As such, the 21,600 points to be clustered exist on a (*P* − 1)-dimensional spherical manifold, with *P* = 90 being the number of regions in the specified AAL atlas. The cluster centroids should be estimated respecting this geometry, which is not the case with Euclidean k-means (Supplementary Figure S1). Additionally, diametrical clustering acknowledges the antipodal symmetry along both directions of a given eigenvector. The classical LEiDA approach seemingly addresses this axial symmetry by flipping the 90-dimensional leading eigenvector for every time point, *t*, if the number of positive elements exceeds the number of negative elements. Especially in the case of an eigenvector with similar numbers of positive and negative loadings, slight variations can result in sign flips that place otherwise similar eigenvectors in different areas of this region space, which can affect clustering results when antipodal symmetry is not considered (see Supplementary Figure S1). More recent approaches to address this include estimating the cosine distance metric and the use of “k-medoids”, which labels specific observed data points as centroids (Farinha et al., 2021). However, this leaves unresolved the limitation of the sign-flip procedure. By acknowledging that the points are distributed on an antipodally symmetric unit hypersphere using diametrical clustering, we obviate the need for eigenvector sign flips. Supplementary Table S12 highlights that although these two strategies can and do produce convergent centroids in some circumstances, there are instances where the two methods diverge (e.g., see State 6 in Supplementary Figure S11). We view diametrical clustering as a technically more appropriate method for clustering eigenvectors since it explicitly models vectors with unit length and arbitrary sign. Therefore, we suggest it is used in future studies investigating dFC using LEiDA.

In this study, we did not address the question of the optimal number of brain states (i.e., cluster centroids). Rather, we explored a range of *k*, 2 to 20, consistent with previous studies (Cabral et al., 2017; Figueroa et al., 2019; Kringelbach et al., 2020). An encouraging sign of the robustness of our observations is that cluster centroids were robust to initialization (Supplementary Figure S10) and stable across *k* (Supplementary Figure S9). Opportunities remain for developing the methodology surrounding the clustering of dynamic BOLD time series. Like other k-means methods, diametrical clustering applies a hard class assignment. Probabilistic estimates of cluster assignment for points on a spherical manifold can be estimated using the Watson mixture model, and non-circular cluster outlines can be estimated using the Bingham distribution (Bingham, 1974; Sra and Karp, 2013; Watson, 1965). These methods are not commonly used, and their development may assist in clustering dynamic functional connectivity data structures and more objectively estimating how many brain states to include. Finally, retaining only the first eigenvector from the eigenvalue decomposition of the phase coherence matrix may remove meaningful information. The rank of a matrix with cosine entries is always two since the angle difference identity allows us to construct two linearly independent vectors, cosine and sine of the input vector, respectively, that fully characterize all information in the input matrix. The instantaneous leading eigenvector constructed as part of the LEiDA pipeline is thus some linear combination of those two trigonometric identities. We observed that, on average, 58% of the variance was explained by the first eigenvector. Future methodological studies should consider whether modeling a multivariate Hilbert phase series without explicitly computing coherence maps and their eigenvectors is possible.

We have previously reported a negative association between static functional connectivity within a priori defined resting-state networks, as well as clusters of brain regions showing increased global functional connectivity as a function of PPL and SDI using rs-fMRI data evaluated here (Madsen et al., 2021). Although the orientation of those and the current findings are conceptually convergent, there are differences. The brain states resolved here by our clustering method are not easily translated to canonical resting-state networks. The frontoparietal states observed here share some regional overlap with default mode network elements (blue nodes; precuneus, posterior cingulate cortex, and to some extent ventromedial prefrontal cortex) and executive control network (red nodes; lateral anterior prefrontal cortex, posterior parietal cortex), but notable regions are absent, such as the angular gyrus from the default mode network. Further, some areas related to visual processing are encompassed by the frontoparietal states (blue nodes; lingual gyrus, calcarine sulcus, cuneus). Together, our studies present complementary perspectives on the associations between resting-state connectivity and PPL and SDI.

The current study examined only acute psilocybin effects on dynamic functional connectivity, while previous studies indicate that psilocybin induces lasting changes in clinical symptoms, mood, and core personality traits. To date, three studies have examined long-term psilocybin effects on functional brain imaging, both primarily examining static connectivity measures (Barrett et al., 2020; Doss et al., 2021; McCulloch et al., 2021b). Two of these studies analyzed dynamic conditional correlation (Engle, 2002) as a variance measure of edge-specific correlation coefficients (Barrett et al., 2020; Doss et al., 2021). Further evaluation of lasting modulation of connectivity dynamics will provide complementary insight into the neurobiological mechanisms underlying lasting behavioral and clinical effects of psilocybin.

Our model estimates that a plasma psilocin level of 20 µg/L, corresponding to 70% neocortex 5-HT2AR occupancy (Madsen et al., 2019), results in a more than 50% decrease in the fractional occurrence of frontoparietal state 1 (for *k* = 7). Although this indicates a pronounced change in this brain state, the fact that two participants showed lower fractional occurrence values at baseline indicates individual variability in these connectivity motifs that needs to be understood more thoroughly. The absence of a brain state identifiable only before or after drug administration suggests that even marginal changes in connectivity dynamics may encompass profound perceptual alterations induced by psychedelics. It is likely that alternative methods for measuring or quantifying functional connectivity dynamics or brain function will provide complementary insights into the neural mechanisms underlying psychedelics. For example, a magnetoencephalography study reported pronounced alterations in resting-state network activity following psilocybin administration (Muthukumaraswamy et al., 2013). Findings across the field to date suggest that relevant acute neural effects of psychedelics remain to be fully explored.

Our study is not without its limitations. Pulse and breathing rate data were not available, and therefore, we could not directly regress physiological noise from our data. To overcome this limitation, we attempted to model noise sources via the anatomical component correction algorithm (Behzadi et al., 2007). Head motion was more prevalent during brain scans following psilocybin administration (Madsen et al., 2021). This is an inherent challenge to scanning participants during peak periods of the psychedelic experience. We performed image realignment and regressed out motion parameters and their first derivatives. Additionally, we excluded two full scan sessions, where motion artifacts were pervasive. Nevertheless, we cannot preclude motion-related effects on our results. Furthermore, despite intriguing convergent evidence of psilocybin effects on connectivity dynamics, our sample size is small (*N* = 15). Clustering in a high-dimensional space exposes risk to the “curse of dimensionality”, where most points in space are equally far away from each other, which can hinder the performance of clustering strategies. Here we modeled 21,600 points, approximately 12x as many points as used in a previous, related study (Lord et al., 2019). Our convergent findings and the stability of our centroids (Supplementary Figures S9–10) support the validity of our findings. Nevertheless, replication in this emergent field is critical, and thus, our results would greatly benefit from replication in other data sets (McCulloch et al., 2021a).

In conclusion, we report that acute psilocybin-induced modulation of brain connectivity dynamics is significantly associated with PPL and SDI. These findings implicate distributed functional motifs in the acute and possibly lasting effects of this drug. Methodologically, we propose an alternative method for clustering eigenvectors that more closely reflects their spherical manifold and sign ambiguity. We also highlight a number of important and relevant analyses of data-driven brain states, including survival analysis of dwell time and assessment of clustering stability.

## Data Availability

All data produced in the present study are available upon reasonable request to the authors.

## Contributions

**Anders S Olsen**: Methodology, Software, Validation, Formal analysis, Data curation, Writing - original draft, Writing - review & editing, Visualization; **Anders Lykkebo-Valløe**: Methodology, Writing - review & editing; **Brice Ozenne**: Methodology, Software, Formal analysis, **Martin K Madsen**: Conceptualization, Investigation, Data curation, Writing - review & editing, Project administration, Funding acquisition; **Dea S Stenbæk**: Conceptualization, Investigation, Writing - review & editing, Project administration; **Sophia Armand**: Investigation, Writing - review & editing; **Morten Mørup**: Methodology, Writing - review & editing; **Melanie Ganz**: Methodology, Writing - review & editing; **Gitte M Knudsen**: Conceptualization, Resources, Writing - review & editing, Supervision, Funding acquisition; **Patrick M Fisher**: Conceptualization, Methodology, Investigation, Data curation, Writing - original draft, Writing - review & editing, Supervision, Project administration, Funding acquisition.

## Acknowledgements

We gratefully acknowledge the work of MRI assistants; Maja Rou Marstrand-Joergensen and Albin Arvidsson for assisting with data collection; Agnete Dyssegaard and Arafat Nasser for biobank management; Oliver Overgaard Hansen and Vibeke Dam for assistance at psilocybin interventions; Lone Freyr, Gerda Thomsen, Svitlana Olsen, Peter Jensen and Dorthe Givard for technical/administrative assistance; the BAFA laboratory, University of Chemistry and Technology and the National Institute of Mental Health (Prague, CZ) for the production of psilocybin; Glostrup Apotek (Glostrup, DK) for encapsulation (GMP); and Sys Stybe Johansen and Kristian Linnet from the University of Copenhagen Department of Forensic Medicine (Copenhagen, DK) for quantification of plasma psilocin levels.

## Clinical trial registration number

NCT03289949, *first registered* 14/09/2017

## Supplementary material

**Supplementary Figure S1:**
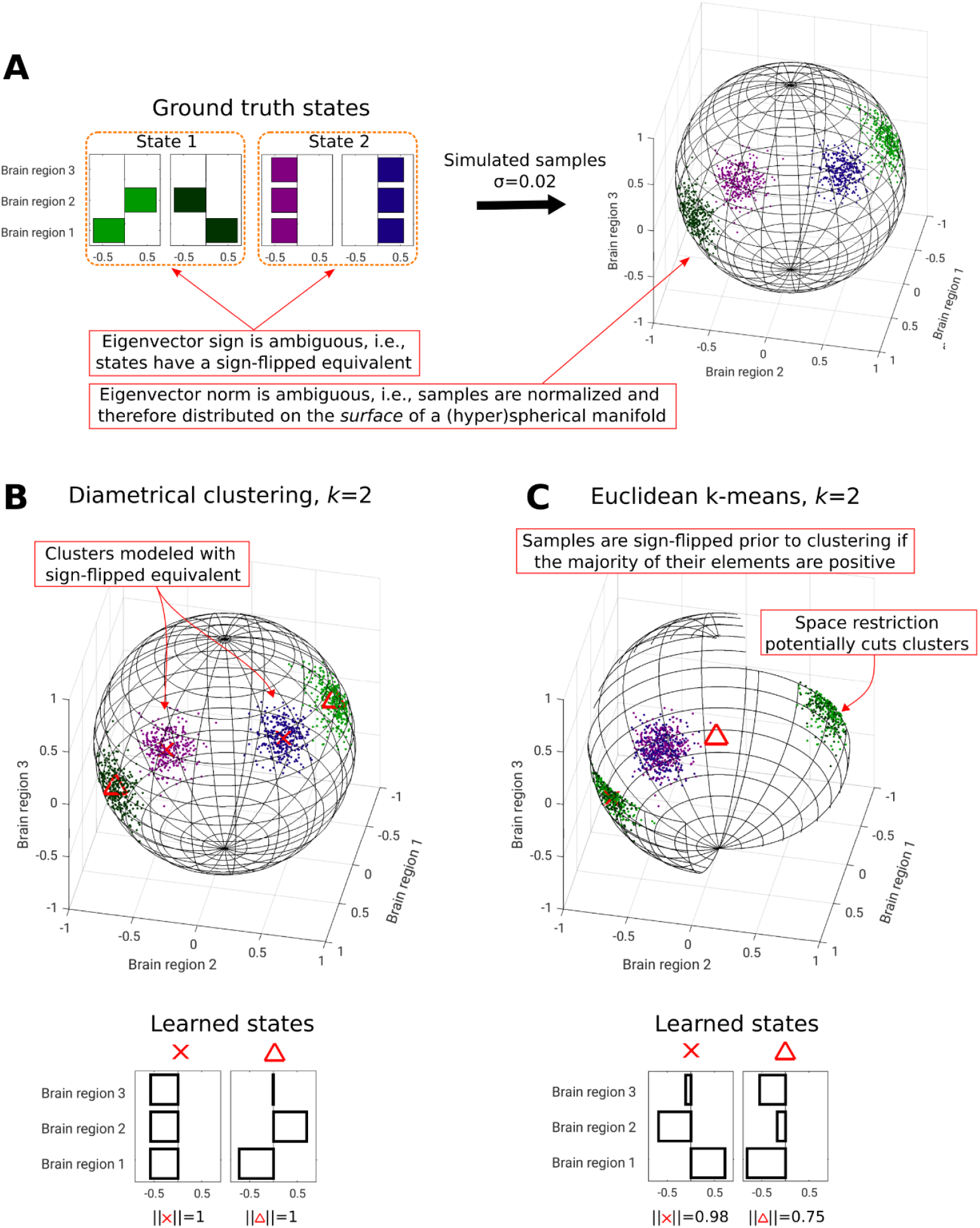
Three-dimensional illustration highlighting the distribution of eigenvectors on an antipodally symmetric unit (hyper)sphere (A), and the conceptual advantage of clustering with respect to this manifold (B) over Euclidean distance clustering (C), which is susceptible to locating centroids off the manifold, where observed points cannot exist. In LEiDA, a sign-flip procedure is applied before k-means clustering, which restricts observation space, potentially cutting through data clusters. The learned states for Euclidean k-means are inferior to diametrical clustering in this hypothetical example.

*Supplementary Video S2: Please find the video here: https://xtra.nru.dk/downloads/misc/AllCentroids_psilocybinDFC.avi. Estimated brain states using LEiDA and diametrical clustering for number of clusters, k, in the range 2 to 20. For k ≥ 3, states were ordered by cycling through the estimated brain states for k − 1 and, for each brain state in the previous k, selecting the brain state for the current k with the largest squared Pearson correlation coefficient. If this coefficient was negative, the new brain state was flipped for visualization purposes. We highlight three brain states: The fully connected (FC) state for all k, the frontoparietal state 1 (FP1) for k ≥ 4, and FP2 for k ≥ 8.*

**Supplementary Table S3:**
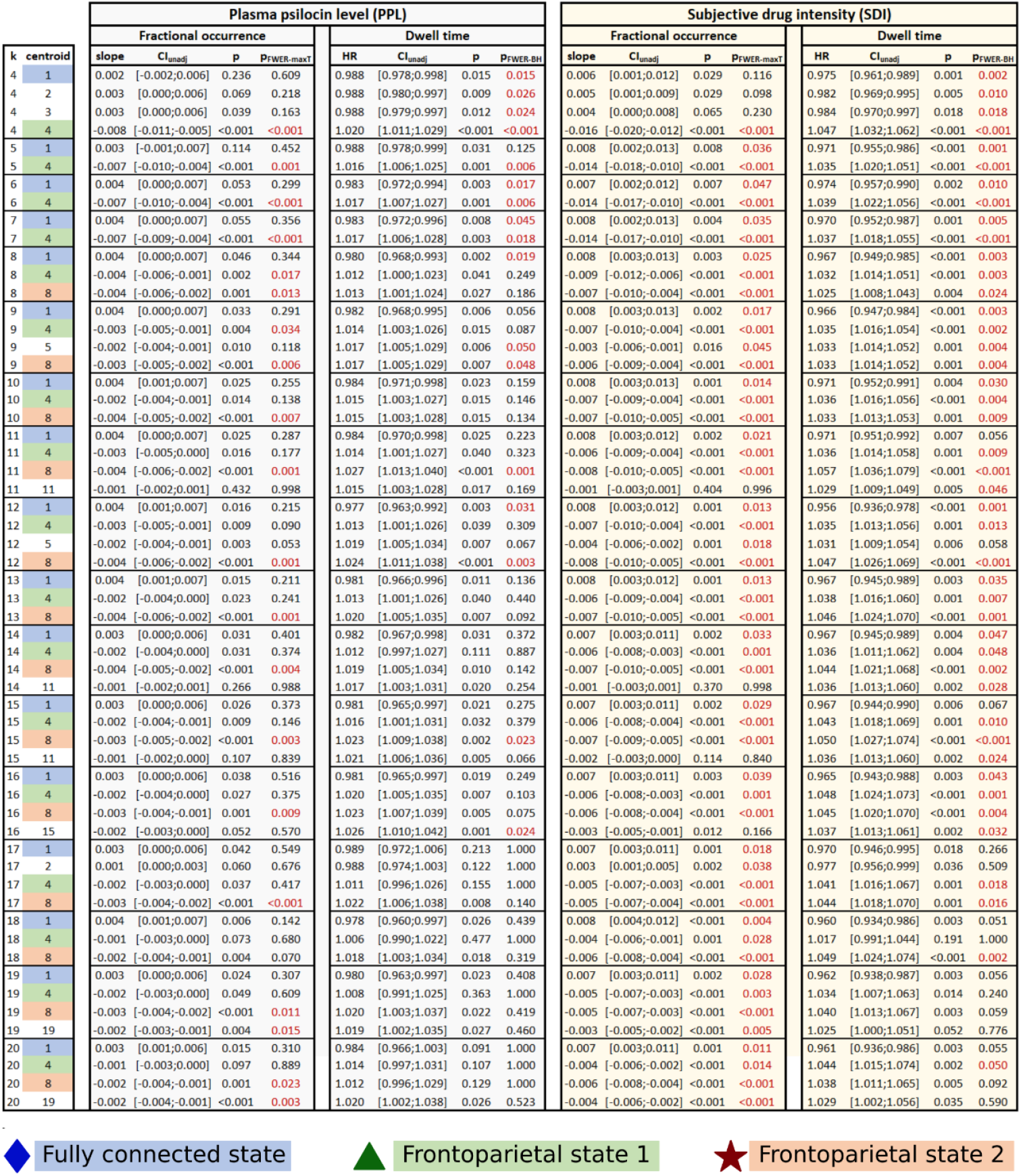
Summary of statistical associations between either fractional occurrence or dwell time and PPL or SDI. Statistical parameters are shown for all brain states for which at least one of the four statistical models was statistically significant after FWER-correction, i.e., either max-T permutation testing for fractional occurrence (p_FWER-maxT_) or Bonferroni-Holm (p_FWER-BH_) for dwell time. Assigned centroid have been ordered consistently with their first appearance (see Supplementary Video S1). As such, the fully connected state is number 1, frontoparietal state 1 is number 4 and frontoparietal state 2 is number 8. p, uncorrected p-value; HR, hazard ratio; CI_unadj_, unadjusted 95% confidence interval.

**Supplementary Figure S4:**
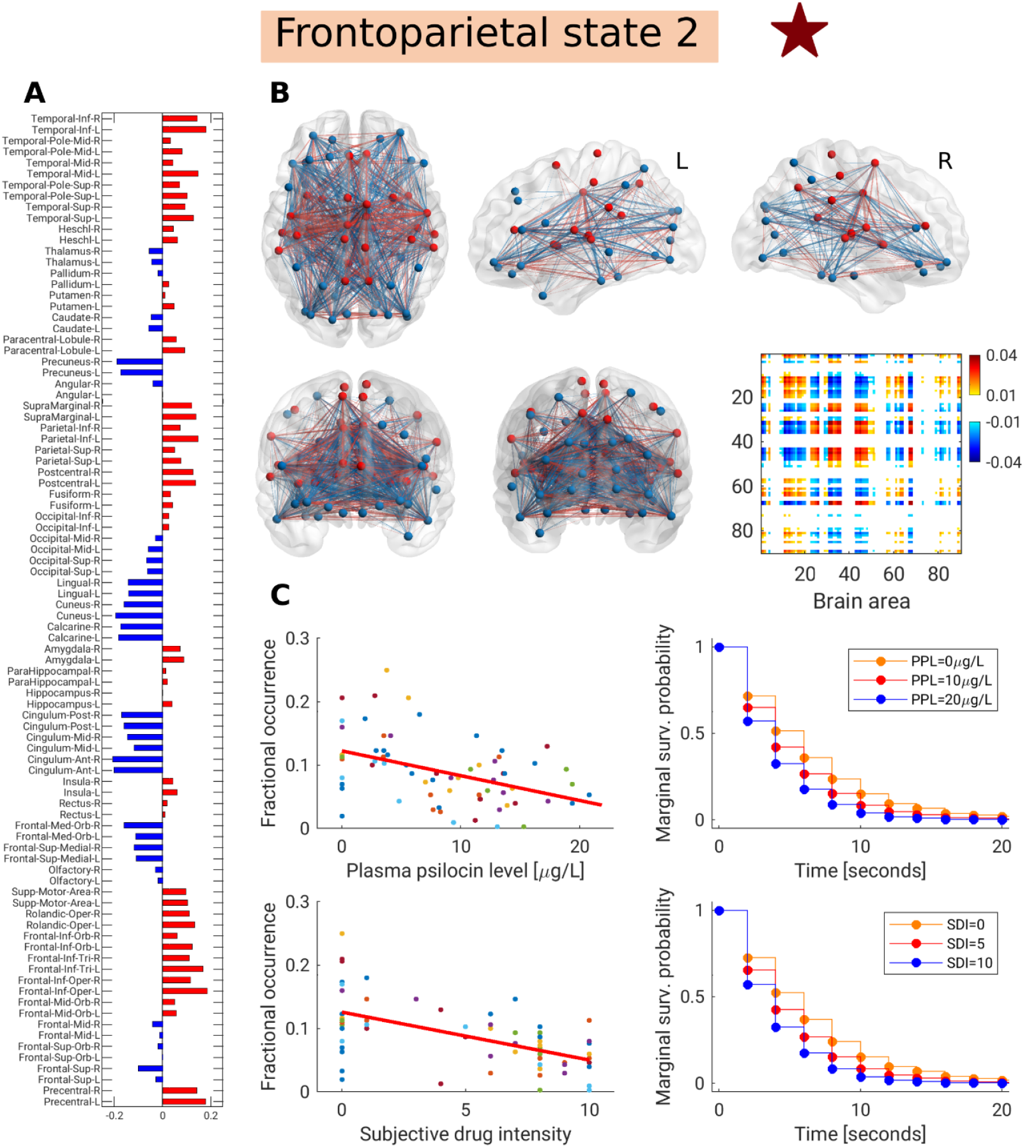
Frontoparietal state 2 and statistical associations for k=11. (A): 90-dimensional centroid, where region pairs with the same sign are said to be coherent. Some frontal regions, superior and inferior parietal regions, and temporal lobe regions showed coherence with each other (red). Likewise, areas around the parieto-occipital sulcus, cingulum, and medial orbital frontal cortex were coherent (blue). (B): Functional coherence map and connectivity representation of the brain state. Edges are shown if their strength exceeded the 75^th^ percentile of absolute edge strengths. In the connectivity visualizations, negative edges are blue and positive edges are red, while nodes are colored according to the sign of their centroid element in (A). (C): Associations between the expression of frontoparietal state 2 and plasma psilocin level and subjective drug intensity using linear mixed models for fractional occurrence (left, each point is a scan session) and frailty Cox proportional hazards models for the dwell time (right). For dwell time, the marginal survival curves for pre-specified covariate levels are shown. Colors in plots of fractional occurrence (C, left) denote individual participants.

**Supplementary Figure S5:**
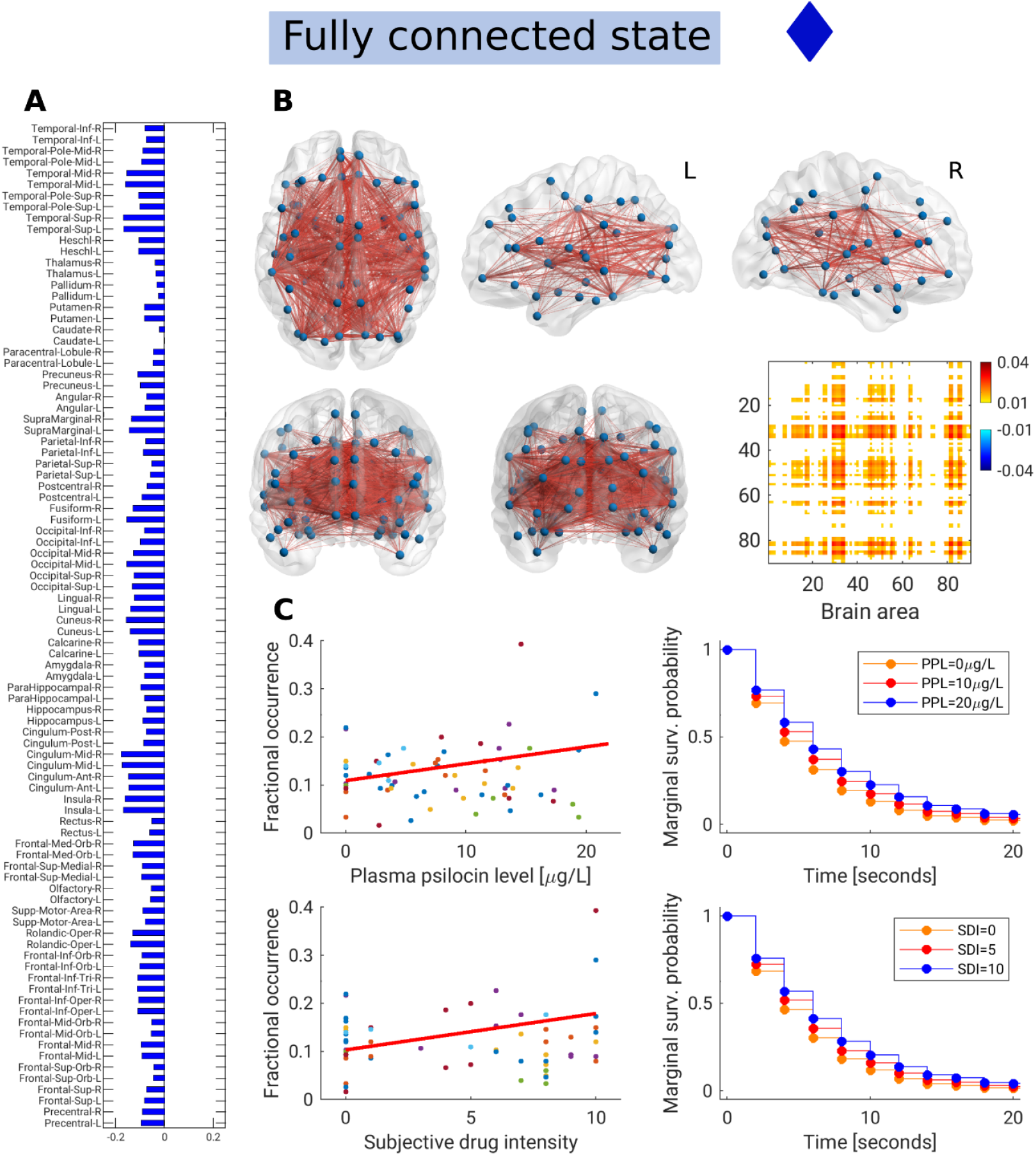
Fully connected state and statistical associations for k=7. (A): 90-dimensional centroid, where region pairs with the same sign are said to be coherent. All centroid loadings for the fully connected state have the same sign, and thus display coherence across all regions. (B): Functional coherence map and connectivity representation of the fully connected state. Edges (red) are shown if their strength exceeded the 75^th^ percentile of absolute edge strengths. In the connectivity visualizations, negative edges are blue and positive edges are red, while nodes are colored according to the sign of their centroid element in (A). (C): Associations between the activity of the full connected state and plasma psilocin level and subjective drug intensity using linear mixed models for fractional occurrence (left, each point is a scan session) and frailty Cox proportional hazards models for the dwell time (right). For dwell time, the marginal survival curves for pre-specified covariate levels are shown. Colors in plots of fractional occurrence (C, left) denote individual participants.

**Supplementary Figure S6:**
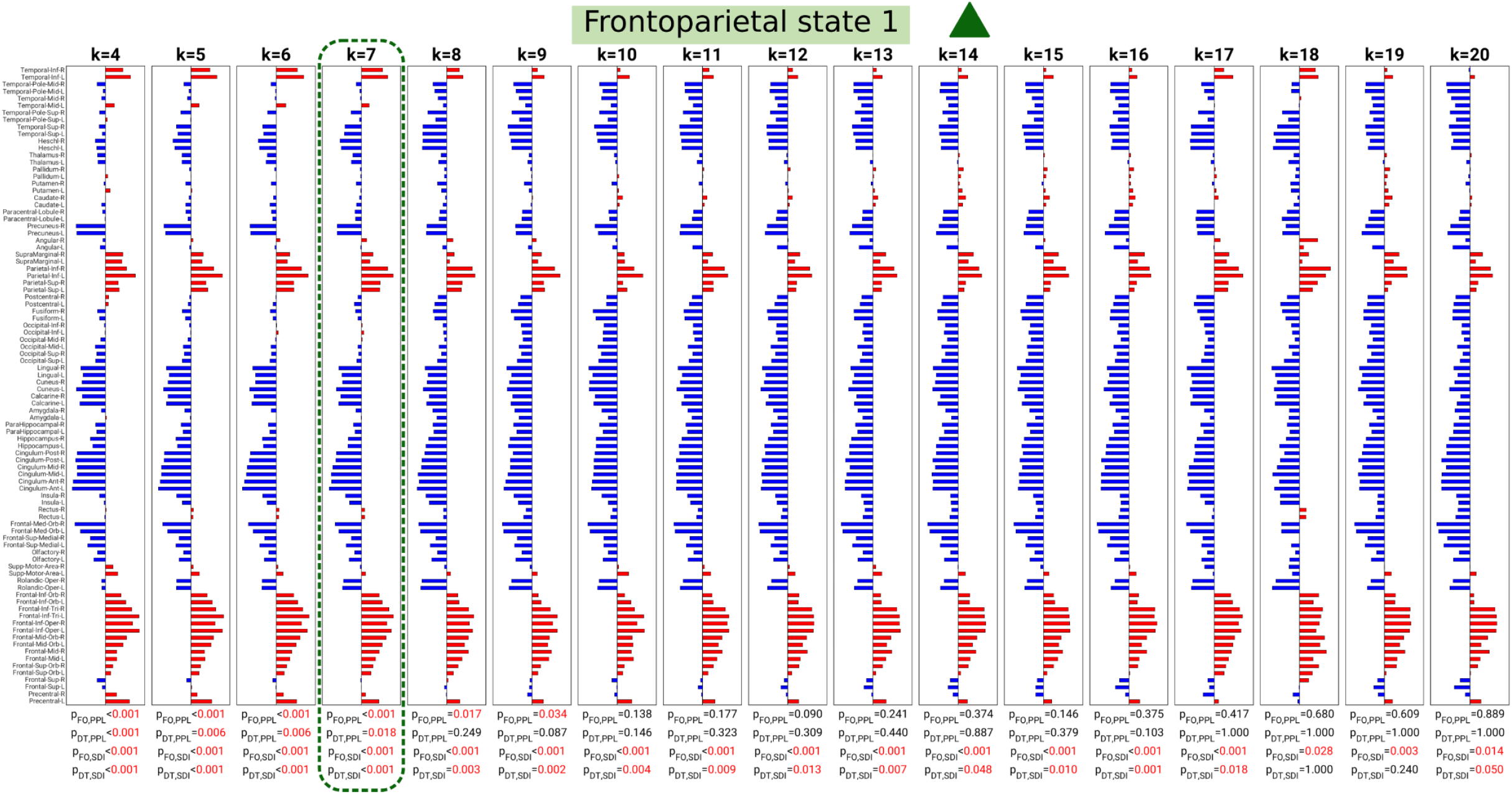
Frontoparietal state 1 centroid across k. For each k ≥ 4, the centroid most similar to the template (highlighted) was selected. P-values at the bottom indicate family-wise error rate corrected statistical significance of the association between either fractional occurrence (FO) or dwell time (DT) and either plasma psilocin level (PPL) or subjective drug intensity (SDI). P-values below 0.05 highlighted in red text.

**Supplementary Figure S7:**
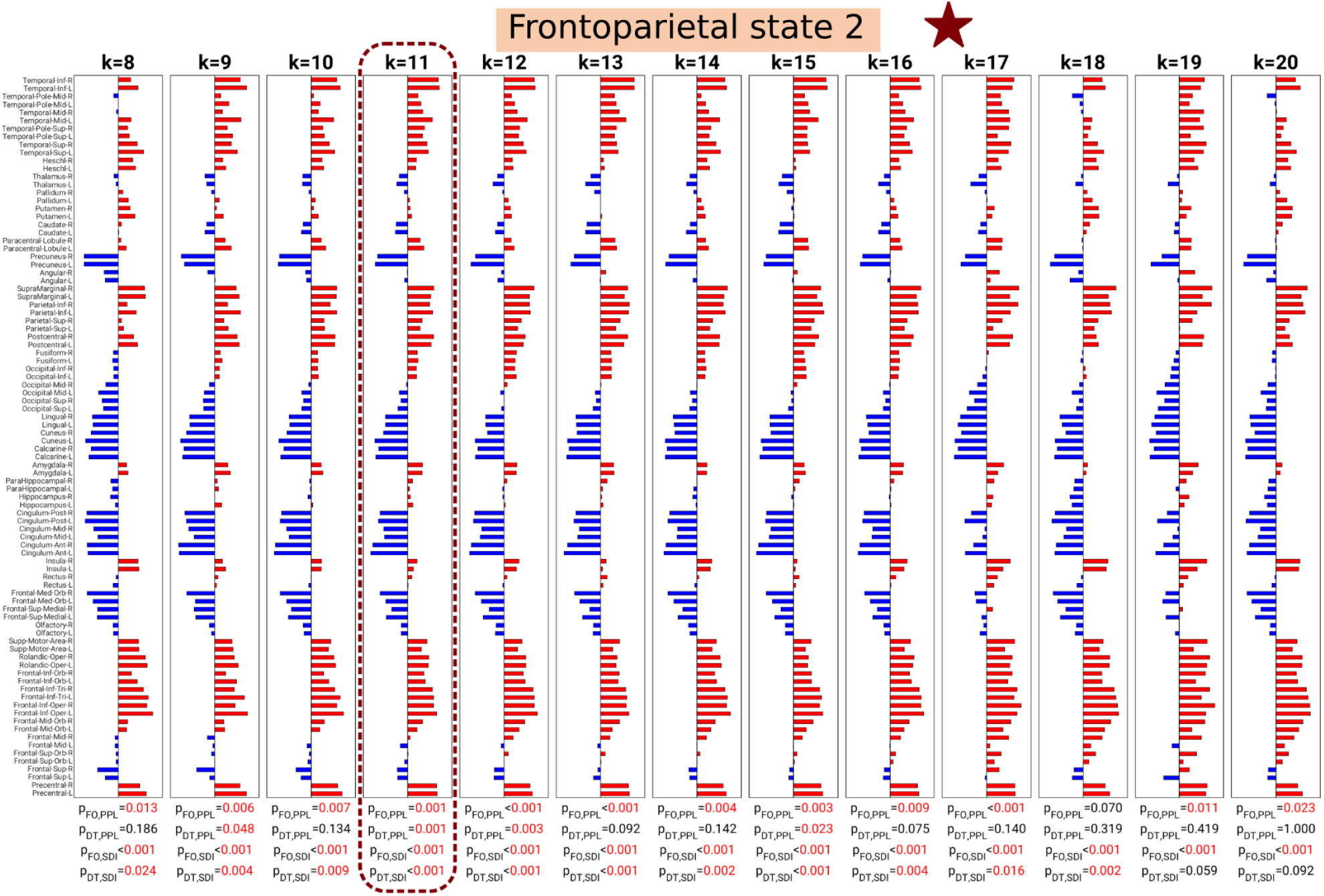
Frontoparietal state 2 centroids across k. For each k ≥ 8, the centroid most similar to the template (highlighted) was selected. P-values at the bottom indicate family-wise error rate corrected statistical significance of the association between either fractional occurrence (FO) or dwell time (DT) and either plasma psilocin level (PPL) or subjective drug intensity (SDI). P-values below 0.05 highlighted in red text.

**Supplementary Figure S8:**
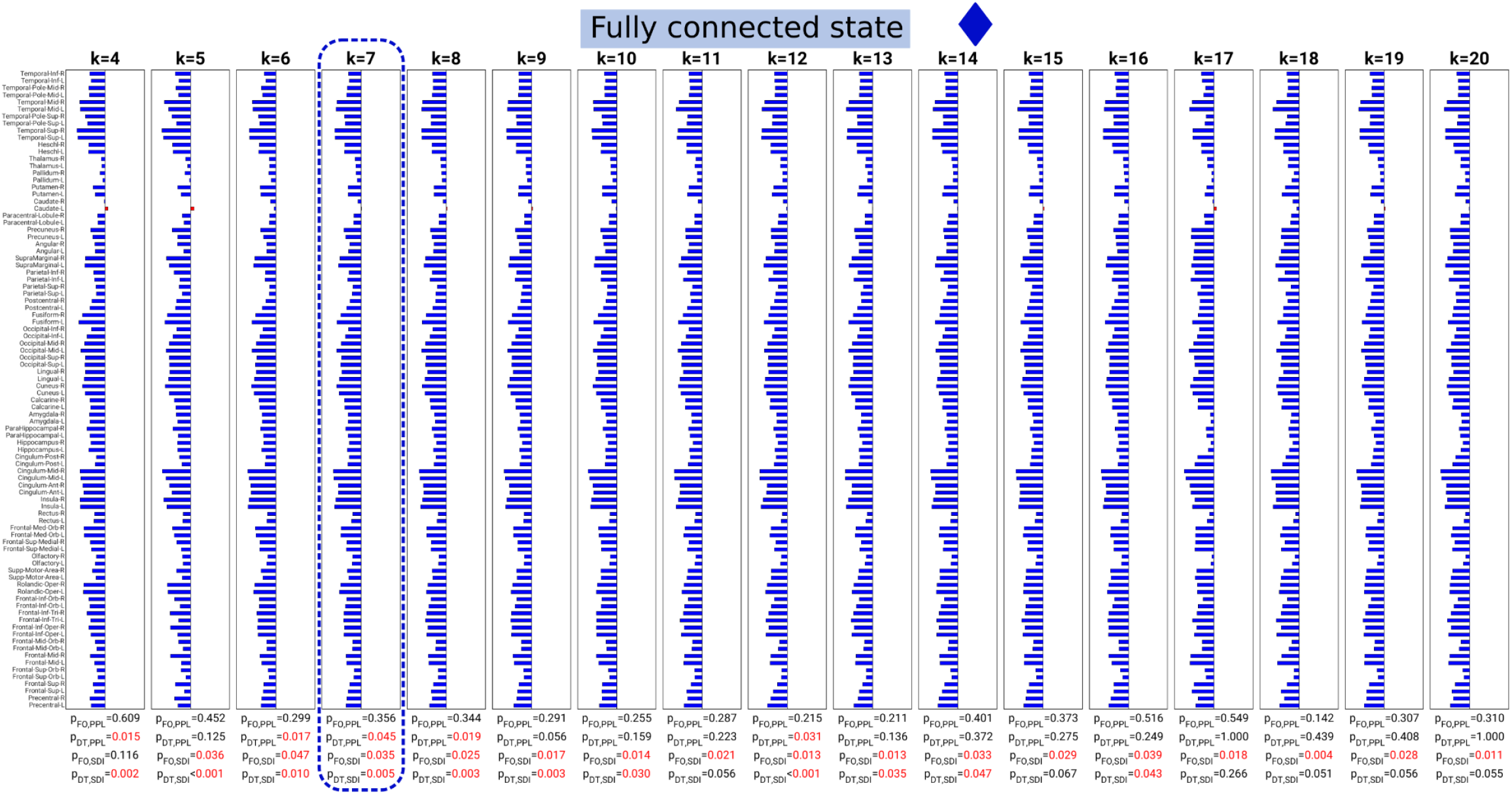
Fully connected state centroids across k. For each k ≥ 4, the centroid most similar to the template (highlighted) was selected. P-values at the bottom indicate family-wise error rate corrected statistical significance of the association between either fractional occurrence (FO) or dwell time (DT) and either plasma psilocin level (PPL) or subjective drug intensity (SDI). P-values below 0.05 highlighted in red text

**Supplementary Figure S9:**
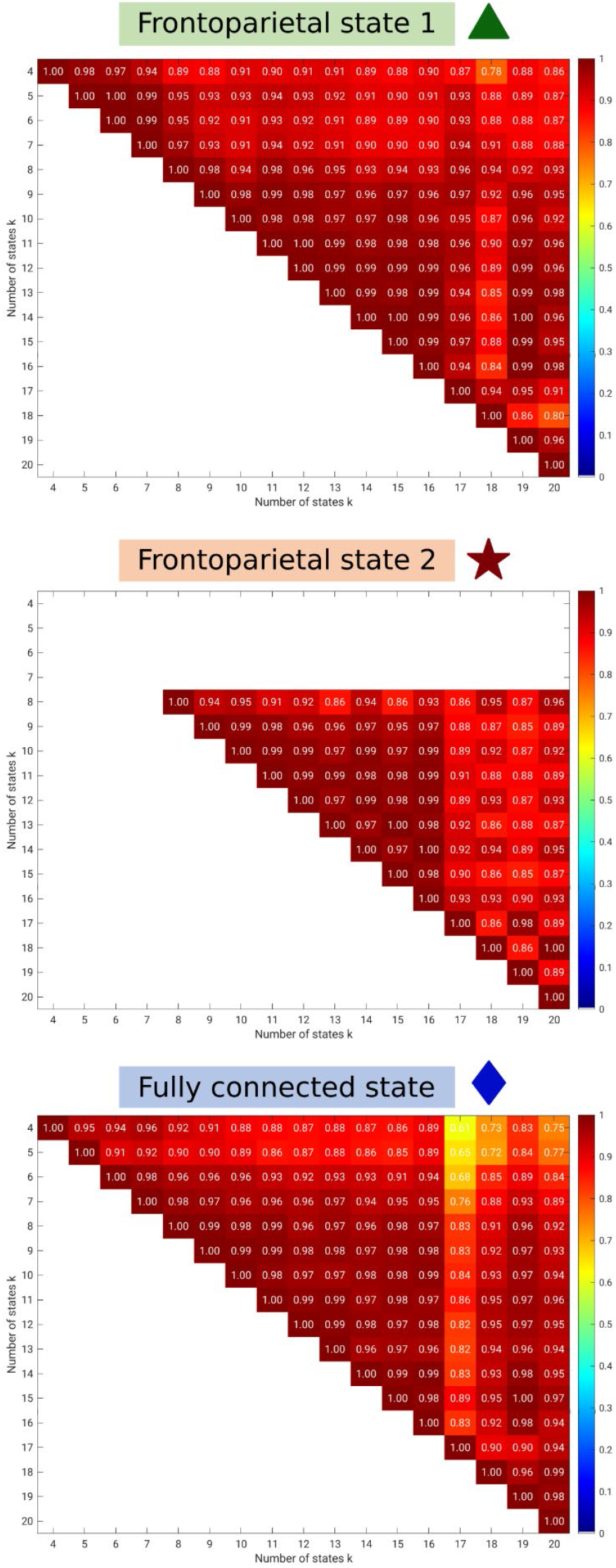
Within-state similarity across k. Heat-map of Pearson correlation coefficients between all pairs of k for each of the three highlighted brain states.

**Supplementary Figure S10:**
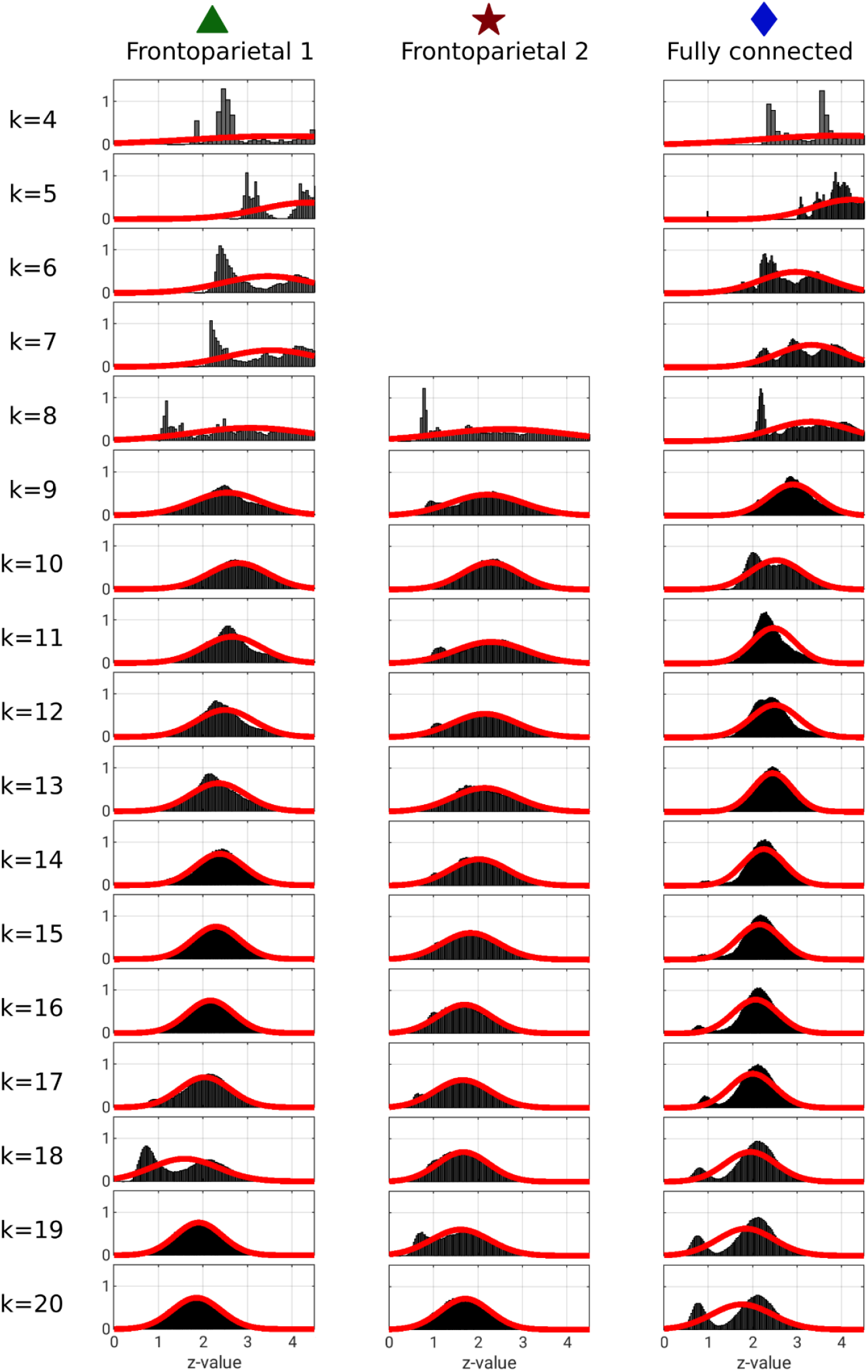
Stability histograms of brain state centroids across 1000 random initializations, each with 5 replications. For every initialization, the relevant centroids were extracted by matching to the three template brain states (see Figures 4 and S4–5). If the identified centroid and template had negative Pearson correlation coefficient, the sign of the identified centroid was flipped. Pearson correlation coefficients between all 1000×1000 brain state pairs in each pool were computed and converted to z-scores using Fisher’s r-to-z transformation, and the corresponding histogram was fitted with a Gaussian curve.

**Supplementary Figure S11:**
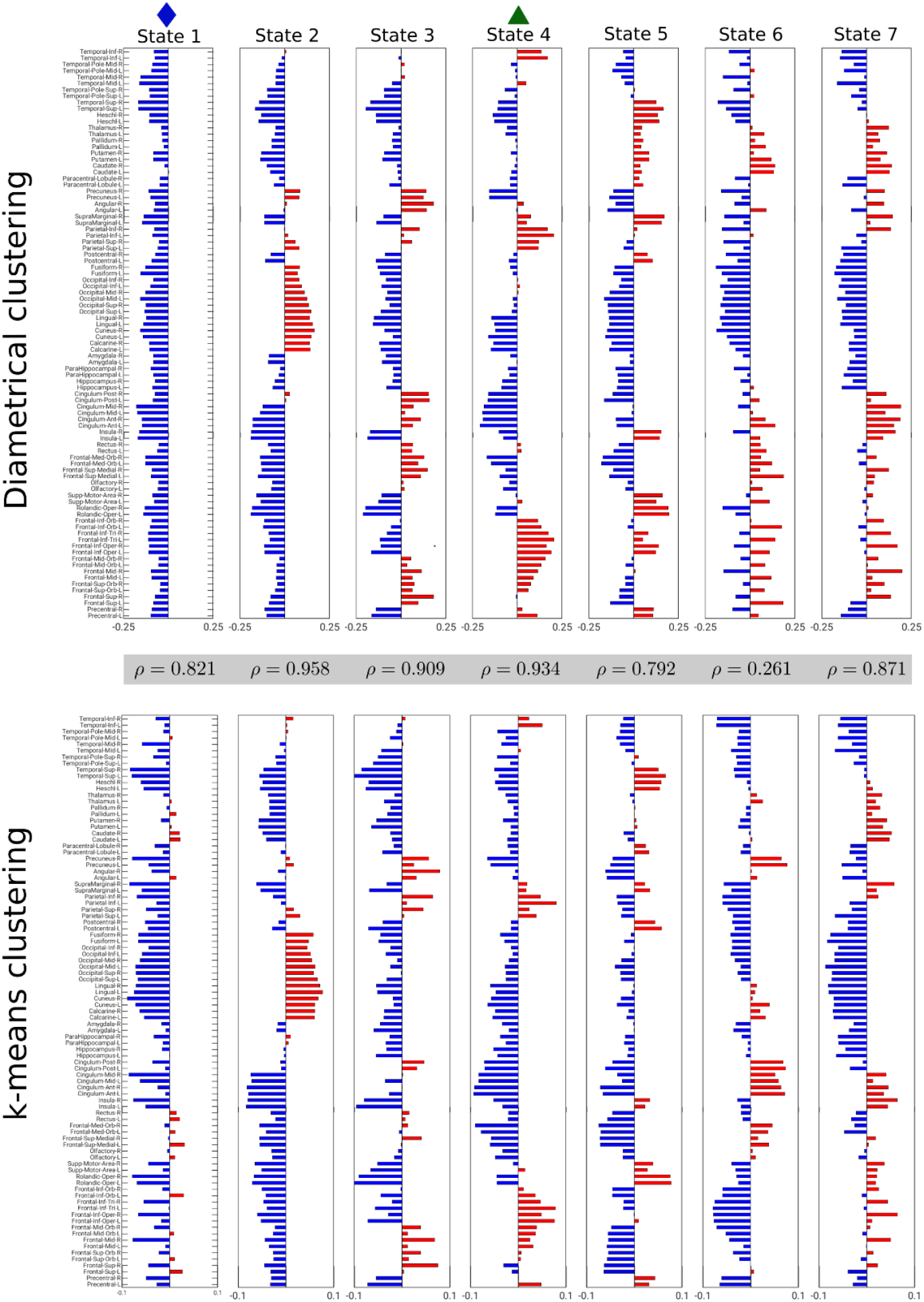
Qualitative differences and correlation coefficients, ρ, between diametrical clustering and Euclidean k-means clustering output centroids at k=7. k-means centroids have been ordered to maximize correlation with diametrical clustering centroids.

**Supplementary Table S12:** Summary Pearson correlation coefficients, ρ, including mean, µ_ρ_, and standard deviation, σ_ρ_, between the highlighted brain states from diametrical clustering and Euclidean k-means clustering. For every k, 1000 initializations of each of the two clustering methods were run, the centroids most closely matching the frontoparietal states 1 and 2 and the fully connected states were extracted, and the Pearson correlation coefficient between all 1000×1000 centroid comparisons computed.

| k | Frontoparietal 1 |  | Frontoparietal 2 |  | Fully connected |  |
| --- | --- | --- | --- | --- | --- | --- |
| | $\mu_\rho$ | $\sigma_\rho$ | $\mu_\rho$ | $\sigma_\rho$ | $\mu_\rho$ | $\sigma_\rho$ |
| 4 | 0.745 | 0.011 |  |  | 0.723 | 0.006 |
| 5 | 0.850 | 0.065 |  |  | 0.727 | 0.041 |
| 6 | 0.901 | 0.090 |  |  | 0.759 | 0.072 |
| 7 | 0.850 | 0.104 |  |  | 0.763 | 0.071 |
| 8 | 0.907 | 0.082 | 0.781 | 0.135 | 0.825 | 0.131 |
| 9 | 0.948 | 0.061 | 0.821 | 0.086 | 0.914 | 0.120 |
| 10 | 0.948 | 0.057 | 0.829 | 0.062 | 0.976 | 0.069 |
| 11 | 0.892 | 0.050 | 0.630 | 0.156 | 0.992 | 0.028 |
| 12 | 0.964 | 0.037 | 0.915 | 0.051 | 0.996 | 0.010 |
| 13 | 0.874 | 0.049 | 0.535 | 0.118 | 0.989 | 0.003 |
| 14 | 0.941 | 0.036 | 0.805 | 0.079 | 0.997 | 0.001 |
| 15 | 0.871 | 0.051 | 0.916 | 0.060 | 0.987 | 0.003 |
| 16 | 0.856 | 0.043 | 0.716 | 0.075 | 0.970 | 0.005 |
| 17 | 0.953 | 0.037 | 0.766 | 0.084 | 0.992 | 0.002 |
| 18 | 0.939 | 0.068 | 0.947 | 0.093 | 0.987 | 0.003 |
| 19 | 0.900 | 0.055 | 0.914 | 0.056 | 0.990 | 0.003 |
| 20 | 0.952 | 0.040 | 0.874 | 0.162 | 0.996 | 0.004 |

## References

Allen, E.A., Damaraju, E., Plis, S.M., Erhardt, E.B., Eichele, T., Calhoun, V.D., 2014. Tracking whole-brain connectivity dynamics in the resting state. Cereb. Cortex 1, 663–676. https://doi.org/10.1093/cercor/bhs352

Arthur, D., Vassilvitskii, S., 2007. K-means++: The advantages of careful seeding, in: Proceedings of the Annual ACM-SIAM Symposium on Discrete Algorithms. pp. 1027–1035.

Barrett, F.S., Doss, M.K., Sepeda, N.D., Pekar, J.J., Griffiths, R.R., 2020. Emotions and brain function are altered up to one month after a single high dose of psilocybin. Sci. Rep. 1, 1–14. https://doi.org/10.1038/s41598-020-59282-y

Bastos, A.M., Schoffelen, J.M., 2016. A tutorial review of functional connectivity analysis methods and their interpretational pitfalls. Front. Syst. Neurosci. 1, 1–23. https://doi.org/10.3389/fnsys.2015.00175

Behzadi, Y., Restom, K., Liau, J., Liu, T.T., 2007. A component based noise correction method (CompCor) for BOLD and perfusion based fMRI. Neuroimage 1, 90–101. https://doi.org/10.1016/j.neuroimage.2007.04.042

Bingham, C., 1974. An Antipodally Symmetric Distribution on the Sphere. Ann. Stat. 1, 1201–1225. https://doi.org/10.1214/aos/1176342874

Bogenschutz, M.P., Forcehimes, A.A., Pommy, J.A., Wilcox, C.E., Barbosa, P., Strassman, R.J., 2015. Psilocybin-assisted treatment for alcohol dependence: A proof-of-concept study. J. Psychopharmacol. 29, 289–299. https://doi.org/10.1177/0269881114565144

Cabral, J., Vidaurre, D., Marques, P., Magalhães, R., Silva Moreira, P., Miguel Soares, J., Deco, G., Sousa, N., Kringelbach, M.L., 2017. Cognitive performance in healthy older adults relates to spontaneous switching between states of functional connectivity during rest. Sci. Rep. 1, 1–13. https://doi.org/10.1038/s41598-017-05425-7

Calhoun, V.D., Miller, R., Pearlson, G., Adali, T., 2014. The Chronnectome: Time-Varying Connectivity Networks as the Next Frontier in fMRI Data Discovery. Neuron 1, 262–274. https://doi.org/10.1016/j.neuron.2014.10.015

Carhart-Harris, R.L., Bolstridge, M., Day, C.M.J., Rucker, J., Watts, R., Erritzoe, D.E., Kaelen, M., Giribaldi, B., Bloomfield, M., Pilling, S., Rickard, J.A., Forbes, B., Feilding, A., Taylor, D., Curran, H. V., Nutt, D.J., 2018. Psilocybin with psychological support for treatment-resistant depression: six-month follow-up. Psychopharmacology (Berl). 1, 399–408. https://doi.org/10.1007/s00213-017-4771-x

Carhart-Harris, R.L., Giribaldi, B., Watts, R., Baker-Jones, M., Murphy-Beiner, A., Murphy, R., Martell, J., Blemings, A., Erritzoe, D., Nutt, D.J., 2021. Trial of Psilocybin versus Escitalopram for Depression. N. Engl. J. Med. 1, 1402–1411. https://doi.org/10.1056/nejmoa2032994

Cox, D.R., 1972. Regression Models and Life-Tables. J. R. Stat. Soc. Ser. B 1, 187–202. https://doi.org/10.1111/j.2517-6161.1972.tb00899.x

Davis, A.K., Barrett, F.S., May, D.G., Cosimano, M.P., Sepeda, N.D., Johnson, M.W., Finan, P.H., Griffiths, R.R., 2021. Effects of Psilocybin-Assisted Therapy on Major Depressive Disorder: A Randomized Clinical Trial. JAMA Psychiatry 1, 481–489. https://doi.org/10.1001/jamapsychiatry.2020.3285

Deco, G., Kringelbach, M.L., 2016. Metastability and Coherence: Extending the Communication through Coherence Hypothesis Using A Whole-Brain Computational Perspective. Trends Neurosci. 1, 125–135. https://doi.org/10.1016/J.TINS.2016.01.001

Dhillon, I.S., Marcotte, E.M., Roshan, U., 2003. Diametrical clustering for identifying anti-correlated gene clusters. Bioinformatics 1, 1612–1619. https://doi.org/10.1093/bioinformatics/btg209

Doss, M.K., Považan, M., Rosenberg, M.D., Sepeda, N.D., Davis, A.K., Finan, P.H., Smith, G.S., Pekar, J.J., Barker, P.B., Griffiths, R.R., Barrett, F.S., 2021. Psilocybin therapy increases cognitive and neural flexibility in patients with major depressive disorder. Transl. Psychiatry 1, 1–10. https://doi.org/10.1038/s41398-021-01706-y

Engle, R., 2002. Dynamic conditional correlation: A simple class of multivariate generalized autoregressive conditional heteroskedasticity models. J. Bus. Econ. Stat. 1, 339–350. https://doi.org/10.1198/073500102288618487

Erritzoe, D., Roseman, L., Nour, M.M., MacLean, K., Kaelen, M., Nutt, D.J., Carhart-Harris, R.L., 2018. Effects of psilocybin therapy on personality structure. Acta Psychiatr. Scand. 138, 368–378. https://doi.org/10.1111/acps.12904

Escrichs, A., Biarnes, C., Garre-Olmo, J., Fernández-Real, J.M., Ramos, R., Pamplona, R., Brugada, R., Serena, J., Ramió-Torrentà, L., Coll-De-Tuero, G., Gallart, L., Barretina, J., Vilanova, J.C., Mayneris-Perxachs, J., Essig, M., Figley, C.R., Pedraza, S., Puig, J., Deco, G., 2021. Whole-Brain Dynamics in Aging: Disruptions in Functional Connectivity and the Role of the Rich Club. Cereb. Cortex 1, 2466–2481. https://doi.org/10.1093/cercor/bhaa367

Farinha, M., Amado, C., Cabral, J., 2021. Increased excursions to functional networks in schizophrenia in the absence of task. bioRxiv 2021.11.25.469834. https://doi.org/10.1101/2021.11.25.469834

Figueroa, C.A., Cabral, J., Mocking, R.J.T., Rapuano, K.M., van Hartevelt, T.J., Deco, G., Expert, P., Schene, A.H., Kringelbach, M.L., Ruhé, H.G., 2019. Altered ability to access a clinically relevant control network in patients remitted from major depressive disorder. Hum. Brain Mapp. 1, 2771–2786. https://doi.org/10.1002/hbm.24559

Garcia-Romeu, A., Davis, A.K., Erowid, F., Erowid, E., Griffiths, R.R., Johnson, M.W., 2019. Cessation and reduction in alcohol consumption and misuse after psychedelic use. J. Psychopharmacol. 1, 1088–1101. https://doi.org/10.1177/0269881119845793

Glerean, E., Salmi, J., Lahnakoski, J.M., Jääskeläinen, I.P., Sams, M., 2012. Functional Magnetic Resonance Imaging Phase Synchronization as a Measure of Dynamic Functional Connectivity. Brain Connect. 1, 91–101. https://doi.org/10.1089/brain.2011.0068

Griffiths, R.R., Richards, W.A., McCann, U., Jesse, R., 2006. Psilocybin can occasion mystical-type experiences having substantial and sustained personal meaning and spiritual significance. Psychopharmacology (Berl). 1, 268–283. https://doi.org/10.1007/s00213-006-0457-5

Hasler, F., Grimberg, U., Benz, M.A., Huber, T., Vollenweider, F.X., 2004. Acute psychological and physiological affects of psilocybin in healthy humans: A double-blind, placebo-controlled dose-effect study. Psychopharmacology (Berl). 1, 145–156. https://doi.org/10.1007/s00213-003-1640-6

Holm, S., 1979. A Simple Sequentially Rejective Multiple Test Procedure A Simple Sequentially Rejective Multiple Test Procedure. Scand. J. Stat. 1, 65–70.

Kolaczynska, K.E., Liechti, M.E., Duthaler, U., 2021. Development and validation of an LC-MS/MS method for the bioanalysis of psilocybin’s main metabolites, psilocin and 4-hydroxyindole-3-acetic acid, in human plasma. J. Chromatogr. B 1, 122486. https://doi.org/10.1016/J.JCHROMB.2020.122486

Kringelbach, M.L., Cruzat, J., Cabral, J., Knudsen, G.M., Carhart-Harris, R., Whybrow, P.C., Logothetis, N.K., Deco, G., 2020. Dynamic coupling of whole-brain neuronal and neurotransmitter systems. Proc. Natl. Acad. Sci. U. S. A. 1, 9566–9576. https://doi.org/10.1073/pnas.1921475117

Larabi, D.I., Renken, R.J., Cabral, J., Marsman, J.B.C., Aleman, A., Ćurčić-Blake, B., 2020. Trait self-reflectiveness relates to time-varying dynamics of resting state functional connectivity and underlying structural connectomes: Role of the default mode network. Neuroimage 1, 116896. https://doi.org/10.1016/j.neuroimage.2020.116896

Lee, O.E., Braun, T.M., 2012. Permutation Tests for Random Effects in Linear Mixed Models. Biometrics 1, 486–493. https://doi.org/10.1111/j.1541-0420.2011.01675.x

Lord, L.D., Expert, P., Atasoy, S., Roseman, L., Rapuano, K., Lambiotte, R., Nutt, D.J., Deco, G., Carhart-Harris, R.L., Kringelbach, M.L., Cabral, J., 2019. Dynamical exploration of the repertoire of brain networks at rest is modulated by psilocybin. Neuroimage 1, 127–142. https://doi.org/10.1016/j.neuroimage.2019.05.060

Luppi, A.I., Carhart-Harris, R.L., Roseman, L., Pappas, I., Menon, D.K., Stamatakis, E.A., 2021. LSD alters dynamic integration and segregation in the human brain. Neuroimage 1, 117653. https://doi.org/10.1016/j.neuroimage.2020.117653

MacLean, K.A., Johnson, M.W., Griffiths, R.R., 2011. Mystical experiences occasioned by the hallucinogen psilocybin lead to increases in the personality domain of openness. J. Psychopharmacol. 1, 1453–1461. https://doi.org/10.1177/0269881111420188

Madsen, M.K., Fisher, P.M., Burmester, D., Dyssegaard, A., Stenbæk, D.S., Kristiansen, S., Johansen, S.S., Lehel, S., Linnet, K., Svarer, C., Erritzoe, D., Ozenne, B., Knudsen, G.M., 2019. Psychedelic effects of psilocybin correlate with serotonin 2A receptor occupancy and plasma psilocin levels. Neuropsychopharmacology 1, 1328–1334. https://doi.org/10.1038/s41386-019-0324-9

Madsen, M.K., Fisher, P.M.D., Stenbæk, D.S., Kristiansen, S., Burmester, D., Lehel, S., Páleníček, T., Kuchař, M., Svarer, C., Ozenne, B., Knudsen, G.M., 2020. A single psilocybin dose is associated with long-term increased mindfulness, preceded by a proportional change in neocortical 5-HT2A receptor binding. Eur. Neuropsychopharmacol. 1, 71–80. https://doi.org/10.1016/j.euroneuro.2020.02.001

Madsen, M.K., Stenbæk, D.S., Arvidsson, A., Armand, S., Marstrand-Joergensen, M.R., Johansen, S.S., Linnet, K., Ozenne, B., Knudsen, G.M., Fisher, P.M., 2021. Psilocybin-induced changes in brain network integrity and segregation correlate with plasma psilocin level and psychedelic experience. Eur. Neuropsychopharmacol. 1, 121–132. https://doi.org/10.1016/j.euroneuro.2021.06.001

Mardia, K. V., Jupp, P.E., 1999. Directional Statistics, Directional Statistics. John Wiley and Sons Ltd. https://doi.org/10.1002/9780470316979

Mason, N.L., Kuypers, K.P.C., Müller, F., Reckweg, J., Tse, D.H.Y., Toennes, S.W., Hutten, N.R.P.W., Jansen, J.F.A., Stiers, P., Feilding, A., Ramaekers, J.G., 2020. Me, myself, bye: regional alterations in glutamate and the experience of ego dissolution with psilocybin. Neuropsychopharmacology 1, 2003–2011. https://doi.org/10.1038/s41386-020-0718-8

McCulloch, D.E.-W., Knudsen, G.M., Barrett, F.S., Doss, M., Deco, G., Carhart-Harris, R.L., Rosas, F., Preller, K., Ramaekers, J., Mason, N., Müller, F., Fisher, P.M., 2021a. Psychedelic resting-state neuroimaging: a review and perspective on balancing replication and novel analyses. PsyArXiv 1–30. https://doi.org/10.31234/OSF.IO/64KYG

McCulloch, D.E.-W., Madsen, M.K., Stenbæk, D.S., Kristiansen, S., Ozenne, B., Jensen, P.S., Knudsen, G.M., Fisher, P.M., 2021b. Lasting effects of a single psilocybin dose on resting-state functional connectivity in healthy individuals. J. Psychopharmacol. 026988112110264. https://doi.org/10.1177/02698811211026454

Ml, K. J, Cruzat, J, Cabral, GM K., r, C.-H., PC, W., NK, L., G, D., 2020. Dynamic coupling of whole-brain neuronal and neurotransmitter systems. Proc. Natl. Acad. Sci. U. S. A. 117, 9566–9576. https://doi.org/10.1073/PNAS.1921475117

Muthukumaraswamy, S.D., Carhart-Harris, R.L., Moran, R.J., Brookes, M.J., Williams, T.M., Errtizoe, D., Sessa, B., Papadopoulos, A., Bolstridge, M., Singh, K.D., Feilding, A., Friston, K.J., Nutt, D.J., 2013. Broadband cortical desynchronization underlies the human psychedelic state. J. Neurosci. 1, 15171–15183. https://doi.org/10.1523/JNEUROSCI.2063-13.2013

Preller, K.H., Duerler, P., Burt, J.B., Ji, J.L., Adkinson, B., Stämpfli, P., Seifritz, E., Repovš, G., Krystal, J.H., Murray, J.D., Anticevic, A., Vollenweider, F.X., 2020. Psilocybin Induces Time-Dependent Changes in Global Functional Connectivity. Biol. Psychiatry 1, 197–207. https://doi.org/10.1016/j.biopsych.2019.12.027

Preller, K.H., Razi, A., Zeidman, P., Stämpfli, P., Friston, K.J., Vollenweider, F.X., 2019. Effective connectivity changes in LSD-induced altered states of consciousness in humans. Proc. Natl. Acad. Sci. U. S. A. 1, 2743–2748. https://doi.org/10.1073/pnas.1815129116

Preti, M.G., Bolton, T.A., Van De Ville, D., 2017. The dynamic functional connectome: State-of-the-art and perspectives. Neuroimage 1, 41–54. https://doi.org/10.1016/j.neuroimage.2016.12.061

Roseman, L., Leech, R., Feilding, A., Nutt, D.J., Carhart-Harris, R.L., 2014. The effects of psilocybin and MDMA on between-network resting state functional connectivity in healthy volunteers. Front. Hum. Neurosci. 8. https://doi.org/10.3389/fnhum.2014.00204

Schindler, E.A.D., Sewell, R.A., Gottschalk, C.H., Luddy, C., Flynn, L.T., Lindsey, H., Pittman, B.P., Cozzi, N. V., D’Souza, D.C., 2021. Exploratory Controlled Study of the Migraine-Suppressing Effects of Psilocybin. Neurotherapeutics 1, 534–543. https://doi.org/10.1007/s13311-020-00962-y

Sewell, R.A., Halpern, J.H., Pope, H.G., 2006. Response of cluster headache to psilocybin and LSD. Neurology 1, 1920–1922. https://doi.org/10.1212/01.wnl.0000219761.05466.43

Sra, S., Karp, D., 2013. The multivariate watson distribution: Maximum-likelihood estimation and other aspects. J. Multivar. Anal. 1, 256–269. https://doi.org/10.1016/j.jmva.2012.08.010

Stark, E.A., Cabral, J., Riem, M.M.E., van IJzendoorn, M.H., Stein, A., Kringelbach, M.L., 2021. The power of smiling: The adult brain networks underlying learned infant emotionality. Cereb. Cortex 1, 2019–2029. https://doi.org/10.1093/CERCOR/BHZ219

Stenbæk, D.S., Madsen, M.K., Ozenne, B., Kristiansen, S., Burmester, D., Erritzoe, D., Knudsen, G.M., Fisher, P.M.D., 2021. Brain serotonin 2A receptor binding predicts subjective temporal and mystical effects of psilocybin in healthy humans. J. Psychopharmacol. 1, 459–468. https://doi.org/10.1177/0269881120959609

Tagliazucchi, E., Carhart-Harris, R., Leech, R., Nutt, D., Chialvo, D.R., 2014. Enhanced repertoire of brain dynamical states during the psychedelic experience. Hum. Brain Mapp. 1, 5442–5456. https://doi.org/10.1002/hbm.22562

Tzourio-Mazoyer, N., Landeau, B., Papathanassiou, D., Crivello, F., Etard, O., Delcroix, N., Mazoyer, B., Joliot, M., 2002. Automated anatomical labeling of activations in SPM using a macroscopic anatomical parcellation of the MNI MRI single-subject brain. Neuroimage 1, 273–289. https://doi.org/10.1006/nimg.2001.0978

Vargas, A.S., Luís, Â., Barroso, M., Gallardo, E., Pereira, L., 2020. Psilocybin as a new approach to treat depression and anxiety in the context of life-threatening diseases-a systematic review and meta-analysis of clinical trials. Biomedicines. https://doi.org/10.3390/biomedicines8090331

Vaupel, J.W., Manton, K.G., Stallard, E., 1979. The impact of heterogeneity in individual frailty on the dynamics of mortality. Demography 1, 439–454. https://doi.org/10.2307/2061224

Vohryzek, J., Deco, G., Cessac, B., Kringelbach, M.L., Cabral, J., 2020. Ghost Attractors in Spontaneous Brain Activity: Recurrent Excursions Into Functionally-Relevant BOLD Phase-Locking States. Front. Syst. Neurosci. 1, 20. https://doi.org/10.3389/fnsys.2020.00020

Vollenweider, F.X., Preller, K.H., 2020. Psychedelic drugs: neurobiology and potential for treatment of psychiatric disorders. Nat. Rev. Neurosci. 1, 611–624. https://doi.org/10.1038/s41583-020-0367-2

Watson, G.S., 1965. Equatorial Distributions on a Sphere. Biometrika 1, 193. https://doi.org/10.2307/2333824

Westfall, P., Young, S., 1993. Resampling-based multiple testing: Examples and methods for p-value adjustment.

Whitfield-Gabrieli, S., Nieto-Castanon, A., 2012. Conn: A Functional Connectivity Toolbox for Correlated and Anticorrelated Brain Networks. Brain Connect. 1, 125–141. https://doi.org/10.1089/brain.2012.0073

Witt, S.T., van Ettinger-Veenstra, H., Salo, T., Riedel, M.C., Laird, A.R., 2021. What Executive Function Network is that? An Image-Based Meta-Analysis of Network Labels. Brain Topogr. 1, 598–607. https://doi.org/10.1007/s10548-021-00847-z

Xia, M., Wang, J., He, Y., 2013. BrainNet Viewer: A Network Visualization Tool for Human Brain Connectomics. PLoS One 1, e68910. https://doi.org/10.1371/journal.pone.0068910

